# Genotype-Phenotype Correlation in RBM10-Associated Syndromes – How Variant Function Shapes a Broad Phenotypic Landscape

**DOI:** 10.1101/2025.08.05.25330579

**Authors:** Jeanne M. V. Bang, Christina R. Fagerberg, Thomas K. Doktor, Mia M. Rosenlund, Santiago M. Lumbreras, Mark Burton, Klaus Brusgaard, Ángel Guerra-Moreno, Sofie Høi, Lenet W. Skovstrøm, Nikolaj A. Nielsen, Qin Hao, Carolina Alves, Lars K. Hansen, Melissa Lees, Pim Suwannarat, Connie Stumpel, Margje Sinnema, Alexander P.A Stegmann, Hilde Van Esch, Chiara De Luca, Christine Van Mol, Andrew Green, Dagmar Wieczorek, Jonathan Rodgers, Julie McGaughran, Veronique Duboc, Khaoula Zaafrane-Khachnaoui, Jill Madden, Pankaj Agrawal, Patrick Rump, Blanca Gener, María Jesús Martínez-González, Jean-Marc Good, Giuseppina Vitiello, Francesco Passaretti, Achille Lolascon, Michael Field, Ellenore M. Martin, Boris Keren, Martine Doco-Fenzy, Tony Yammine, Katharina Steindl, Anita Rauch, Anais Begemann, Gregory Costain, Zhuo Shao, Diana Carli, Giovanni Battista Ferrero, Irene Valenzuela, Marta Codina-Solà, Barbara Masotto, Laura Trujillano, Candy Kumps, Olivier Vanakker, Anand Vasudevan, Maria Rita Passos-Bueno, Erasmo Casella, Fernarnda Bonilla Colomé, Laurence Faivre, Christophe Philippe, Marlin Touma, Lee-Kai Wang, Stanley F. Nelson, Marcello Scala, Vincenzo Nigro, Valeria Capra, Kristen Truxal, Valentina Caceres, Jonathan Levy, Vera Kalscheuer, Andrée Delahaye-Duriez, Juan Valcárcel, Michael Sattler, Brage S. Andresen

**Affiliations:** Department of Biochemistry and Molecular Biology, University of Southern Denmark, Odense M, Denmark; Clinical Genetics, Lillebaelt Hospital, Vejle Hospital, Vejle, Denmark; Department of Clinical Genetics, Odense University Hospital, Odense, Denmark; Department of Regional Health Research, University of Southern Denmark, Denmark; Institute of Clinical Research, University of Southern Denmark, Odense, Denmark; Institute of Structural Biology, Molecular Targets and Therapeutics Center, Helmholtz Munich, Neuherberg Germany; Technical University of Munich, TUM School of Natural Sciences, Bavarian NMR Center and Department of Bioscience, Garching, Germany; Clinical Genome Center, University of Southern Denmark & Region of Southern Denmark, Odense, Denmark; Human Genetics, Department of Clinical Research, University of Southern Denmark, Denmark; Genome Biology Program, Centre for Genomic Regulation, The Barcelona Institute of Science and Technology, Barcelona, Spain; FDNA Inc., Boston, MA, USA; H.C. Andersen Childrens Hospital, Odense University Hospital, Odense, Denmark; North Thames Regional Genetics Service, Great Ormond Street Hospital, London, United Kingdom; Mid-Atlantic Permanente Medical Group, Washington DC, USA; Department of Clinical Genetics, University of Maastricht, The Netherlands; Center for Human Genetics, University Hospitals Leuven, Leuven, Belgium; Human Genetics, Department of Life, Health and Environmental Sciences, University of L’Aquila, L’Aquila, Italy; Department of Pediatrics-Neonatology, St. Augustinusziekenhuis, Wilrijk, Belgium; Department of Clinical Genetics, Children’s Health Ireland, Dublin, Ireland; School of Medicine and Medical Science, University College Dublin, Dublin, Ireland; Institute of Human Genetics, Medical Faculty and University Hospital, Heinrich-Heine University, Düsseldorf, Germany; Genetic Health Queensland, Royal Brisbane and Women’s Hospital, Brisbane, Australia; School of Medicine, The University of Queensland, St Lucia, Brisbane, Australia; The University of Queensland St Lucia, Brisbane, Queensland, Australia; Université Côte d’Azur, Centre Hospitalier Universitaire de Nice, service de genetique medicale, Nice, France; The Manton Center, Boston Children’s Hospital, Harvard Medical School Division of Genetics, Department of Medicine, Boston, USA; Division of Newborn Medicine, Manton Center Gene Discovery Core Division of Genetics and Genomics, Boston Children’s Hospital, Boston, USA; University of Groningen, University Medical Center Groningen, Department of Genetics, Groningen, The Netherlands; Department of Genetics, Cruces University Hospital, Biobizkaia Health Research Institute, Barakaldo, Spain; Department of Pediatrics, Pediatric Neurology Unit, Cruces University Hospital, Pediatric Group, Biobizkaia Health Research Institute, Barakaldo, Spain; Division of Genetic Medicine, Lausanne University Hospital and University of Lausanne, Lausanne, Switzerland; Department of Molecular Medicine and Medical Biotechnology, University of Naples Federico II, Napoli, Italy; Genetics of Learning Disability Service, John Hunter Hospital, Waratah, Australia; Murdoch Children’s Research Institute, Melbourne, Victoria, Sydney, Australia; Department of Genetics, Hôpital Pitié-Salpêtrière, Paris, France; Service de Génétique Chu de Reims, Reims, France; Service de Génétique CHU de Nantes, Nantes, France; Institute of Medical Genetics, University of Zürich, Zürich, Switzerland; Division of Clinical and Metabolic Genetics, The Hospital of Sick Children; Program in Genetics & Genome Biology, SickKids Research Institute, Toronto, Canada; Departments of Pediatrics and Molecular Genetics, University of Toronto, Toronto, Canada; The Genetics Program, North York General Hospital, Toronto, Canada; Department of Pediatrics, University of Toronto, Toronto, Canada; Department of Medical Sciences, University of Torino, Torino, Italy; Department of Clinical and Molecular Genetics, Vall d’Hebron Barcelona Hospital Campus, Vall d’Hebron Hospital Universitari, Barcelona, Spain; Center for Medical Genetics, Ghent University Hospital, Ghent, Belgium; The Royal Women’s Hospital, Victoria, Australia; Department of Genetics and Evolutionary Biology, Institute of Bioscience, São Paulo University, São Paulo, Brazil; Instituto da Criança, Faculdade de Medicina, São Paulo University, São Paulo, Brazil; Department of Child Neurology, Hospital Pequeno Príncipe, Curitiba, Brazil; Inserm UMR1231 GAD, Université de Bourgogne Franche-Comté, Dijon, France; CRMR "Anomalies du Développement et Déficience Intellectuelle", FHU-TRANSLAD, CHU Dijon, Dijon, France; Laboratoire de Génétique, CHR Metz Thionville, Hôpital Mercy, Metz, France; David Geffen School of Medicine, University of California, Los Angeles, USA; Department of Neurosciences, Rehabilitation, Ophthalmology, Genetics, Maternal and Child Health, University of Genoa, Genoa, Italy; Medical Genetics Unit, IRCCS Istituto Giannina Gaslini, Genoa, Italy; Department of Precision Medicine, University of Campania "Luigi Vanvitelli", Naples, Italy; Telethon Institute of Genetics and Medicine, Pozzuoli, Italy; Clinical Genetics and Genomics Unit, IRCCS Institute G. Gaslini, Genoa, Italy; Division of Genetic and Genomic Medicine, Nationwide Children’s Hospital, Columbus, OH, USA; The Genetics Department, Hôpital Universitaire Robert-Debré, Paris, France; Max Planck Institute for Molecular Genetics, Berlin, Germany; Medical genomics and clinical genetics unit, AP-HP, Hôpitaux Universitaires Paris Seine Saint Denis, Hôpital Jean Verdier, Bondy, France

**Author notes:** Corresponding author: Brage S. Andresen. Campusvej 55, 5230 Odense M. Mail. Shared first authorship (Jeanne M. V. Bang, Christina R. Fagerberg).

## Abstract

Severe loss of function variants in the splicing regulatory protein RBM10 are known to cause TARP syndrome, a rare X-linked recessive congenital syndrome. In recent years, individuals with milder phenotypes have been published, suggesting a broader phenotypic spectrum.

We report 37 new individuals with RBM10 variants and compare to 34 published cases. We find that the phenotype can be described as an “RBM10-phenotypic spectrum” which can be further subdivided into two phenotypic groups, TARP syndrome (TARPS) and RBM10 Associated Intellectual Disability (RAID).

Based on phenotype characterizations and functional studies, we describe a clear genotype-phenotype correlation. Splicing analysis of blood samples and CRISPR-edited cells representing different degrees of functional loss of RBM10 demonstrated a pattern of more exon inclusion in response to increased loss of RBM10 function. More inclusion was correlated with increasing phenotype severity. Functional studies of missense variants from the different phenotypic groups confirm this genotype-phenotype correlation and show that different molecular mechanisms can explain the underlying pathological alterations in RBM10 protein function. Interestingly, we show that some missense variants in the RNA binding, RRM2 domain of RBM10 alter RBM10 activity from splicing inhibition to stimulation, likely due to altered RNA binding characteristics.

**Graphical Abstract:** 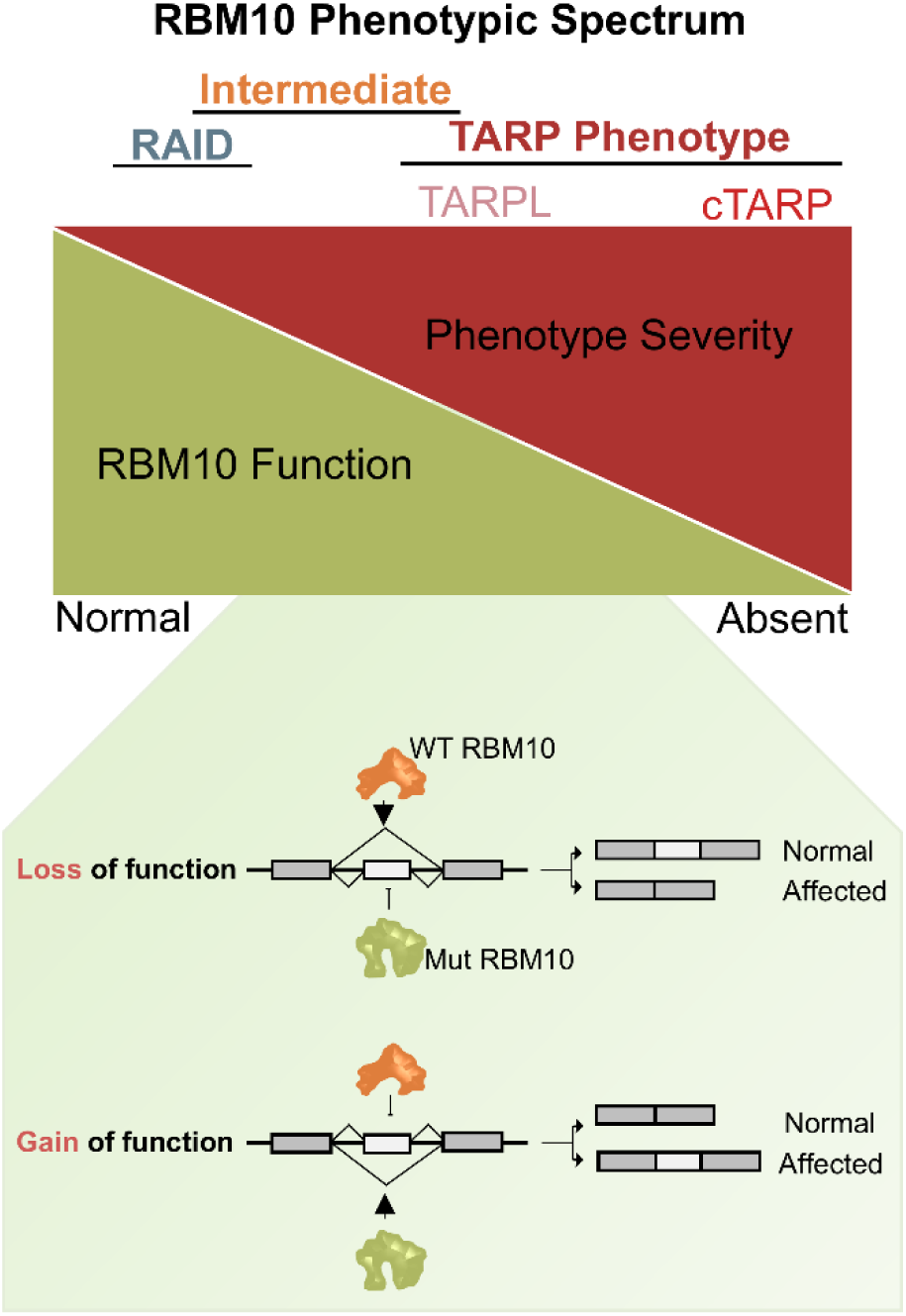

## Introduction

RNA binding motif 10 (*RBM10*) is localized at Xp11.23 (1) and encodes RBM10, a member of the RNA-Binding Motif family of RNA binding proteins (RBP) (2). It is ubiquitously expressed and has multiple roles during normal development through a variety of mechanisms, including splicing regulation, mRNA stabilization, transcription regulation, and apoptosis (2, 3). In addition, RBM10 has been identified as a tumor suppressor in several types of cancer and is also reported to be involved in the development of cardiac hypertrophy (4–6). Functional domains of RBM10 (Figure 1A) include two RNA recognition domains (RRM1 and RRM2), a RanBP2-type zinc finger (ZF) domain, a Cys2-His2-type zinc finger (C_2_H_2_-type ZF) domain, and a Glycine-patch domain (7).

**Figure 1.**
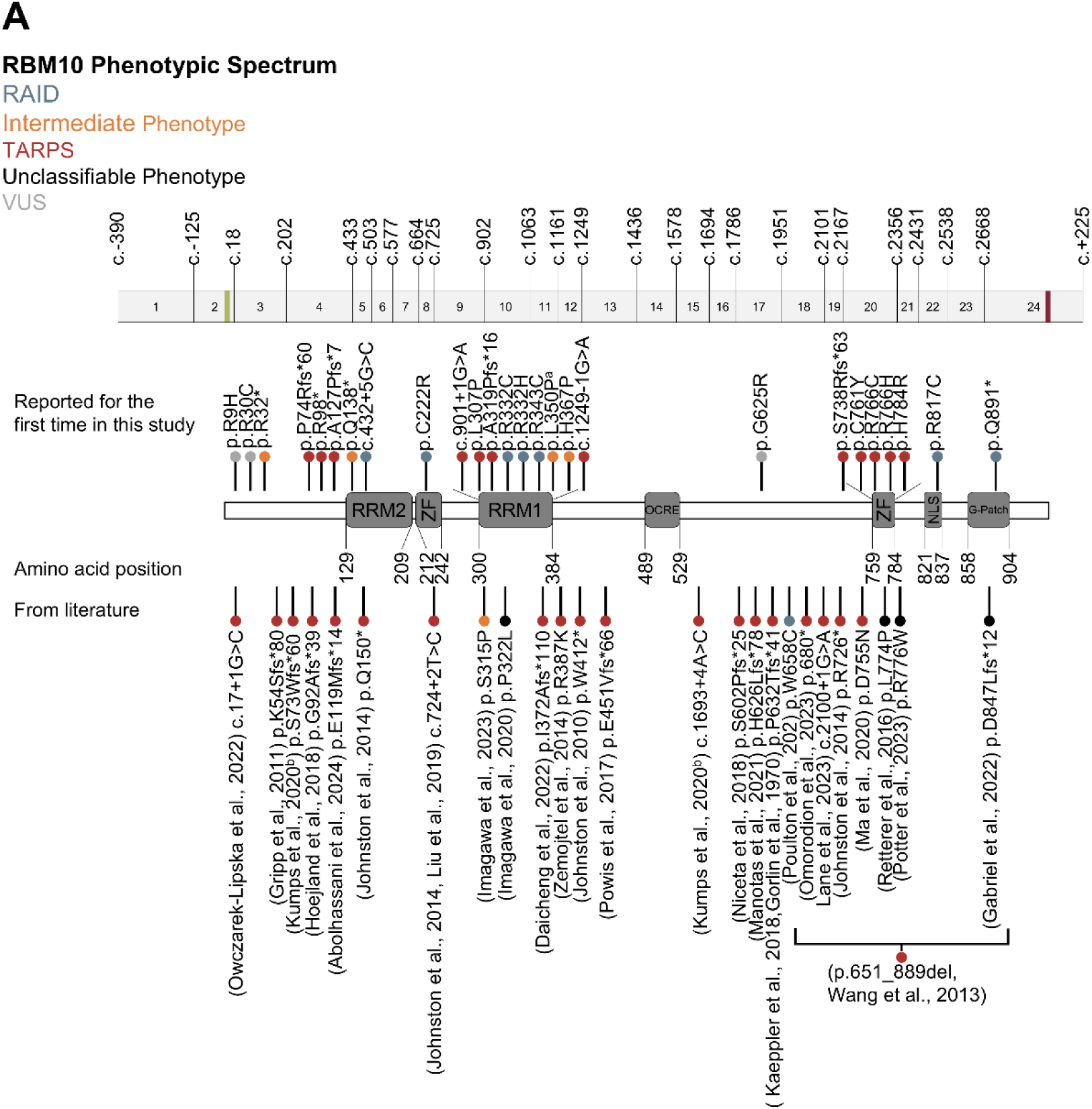
*RBM10* Associated Syndromes: **A.** Schematic Illustration of the *RBM10* gene and protein with exon and domain positions respectively. The functional domains are labelled with amino acid position and exons with cDNA position, green and red mark start and stop codon. Variants in the *RBM10* gene associated with phenotype from the literature are listed under the protein (10, 14–21, 23–34, 36–39). Variants reported for the first time in this study are listed above the protein. NM_005676.5, NP_005667.2. ^a^Individuals included for functional studies. ^b^The p.L350P variant is identified in two siblings, characterized having either RAID (RBM10-associated intellectual disability) or intermediate phenotype. TARPS,(TARP Syndrome), TARPL, (TARP-Like).

*RBM10* undergoes alternative splicing, which results in several RBM10 isoforms. There are two major isoforms: full-length RBM10v1 with all 24 exons included and the shorter RBM10v2 which skips exon 4 (3). RBM10 has a major role in splicing regulation. It is a component of the pre-spliceosomal A and B complex and is associated with several spliceosomal proteins (8, 9). RBM10 is known to regulate splicing by binding to intronic sequences in the vicinity of the splice sites and upstream of the branch point, typically resulting in skipping of cassette exons (CE) which are flanked by weak splice sites (2, 10). RBM10 regulates splicing of *SMN2* exon 7 (11), *NUMB* exon 11 (12), *TNRC6A* exon 7 (7), *BCL2L1* exon 2 (13) and numerous other targets.

Pathogenic loss of function (LOF) variants in the *RBM10* gene cause the X-linked recessive TARP Syndrome (TARPS) (OMIM 311900) (14). TARP is an acronym for **<u>T</u>**alipes Equinovarus, **<u>A</u>**trial septal defect, **<u>R</u>**obin sequence, and **<u>P</u>**ersistent left superior vena cava (15). Most individuals with TARPS however only have one or a few of these features, while the most frequent clinical features are severe global developmental delay, failure to thrive, respiratory insufficiency, hypotonia, brain malformation, and facial dysmorphism (15). While most of the initially reported individuals died in infancy, it has become clear that the phenotype varies much more than initially suspected, and that missense variants can also be disease causing.

So far only three cases with disease causing RBM10 variants have been functionally characterized (10, 16, 17). We characterize a cohort of 37 individuals with *RBM10* variants and compare them to 34 published cases. Moreover, functional characterization of several missense variants reveals a clear genotype-phenotype correlation.

## Results

### Phenotype Grouping

The study includes 71 individuals (all males, 37 from this study (Figure 1A) and 34 published cases (10, 14, 15, 17–33)) with *RBM10* variants. Sixty individuals were included in the comparison of phenotypes (11 individuals were not included because of insufficient phenotypic information (4 from literature (16, 34–36)), because the RBM10-variant was of uncertain significance (5 from our study: P4, P27, P31A, P31B, P31C), or the cases were prenatal (2 from literature (20, 37)) (Supplementary Table S1, familial pedigrees are available upon request). We find that the phenotypes belong to a wide phenotypic spectrum - RBM10-phenotypic spectrum – which can be divided into two major groups: TARPS, a phenotype corresponding to TARP syndrome as originally described, but with variable severity, and RAID, with a different and milder phenotype. Some individuals have an intermediate phenotype with features from TARPS as well as RAID, supporting that these phenotypes do indeed belong to a common spectrum.

### TARP Syndrome, TARPS

Forty-two individuals are categorized as having TARPS, 16 individuals are reported for the first time in this study, and 26 are from literature (10, 14–21, 23–34, 36–39).

TARPS includes individuals with features similar to what was originally described as TARP syndrome. While the phenotype is rather characteristic, the severity of symptoms varies, most importantly concerning survival. To describe the differences in severity within this spectrum, we subdivided TARPS into classical TARP syndrome (cTARP) constituting the most severe end of the spectrum (for practical reasons defined as those with death in infancy, Supplementary Table S2), and TARP-like (TARPL) syndrome, with survival beyond infancy (Table 1). As death in infancy results from different causes and at least partly depends on the available treatment, the border between cTARP and TARPL is however not sharp. Thirteen individuals are classified as having cTARP syndrome, while 29 are classified as having TARPL syndrome.

**Table 1:**
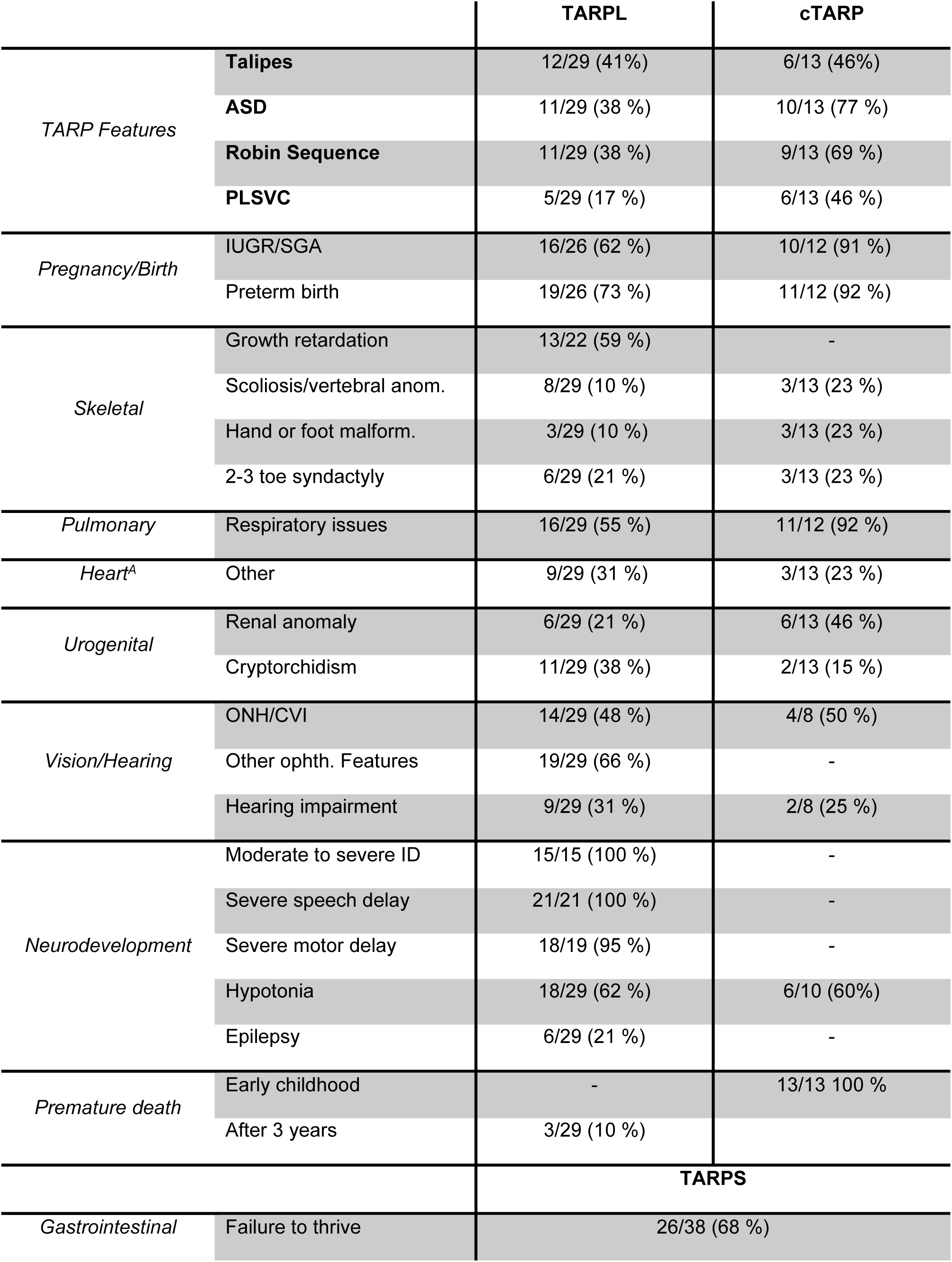
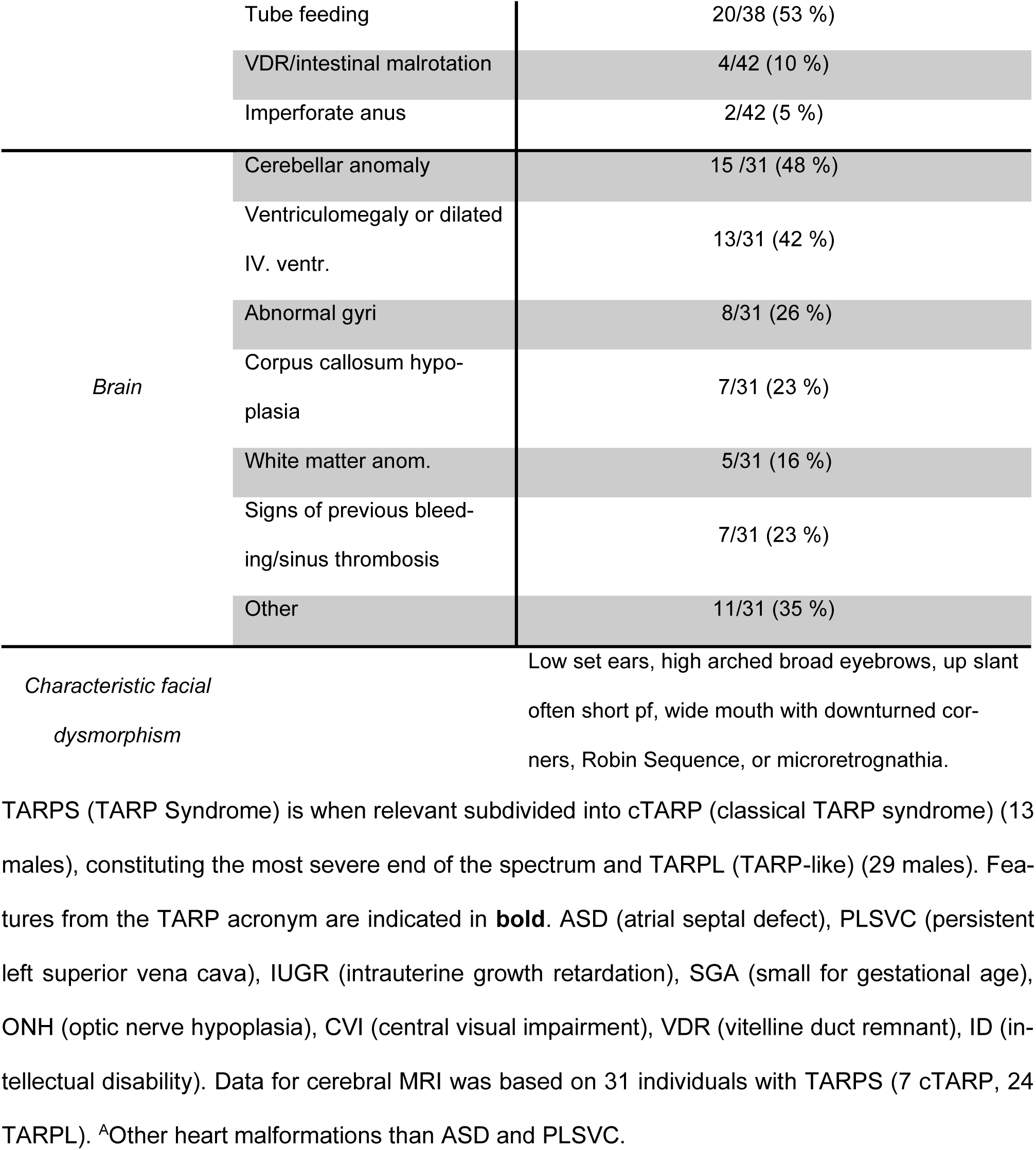
Phenotype of 42 Individuals with TARPS.

|  |  | TARPL | cTARP |
| --- | --- | --- | --- |
| *TARP Features* | **Talipes** | 12/29 (41%) | 6/13 (46%) |
|  | **ASD** | 11/29 (38 %) | 10/13 (77 %) |
|  | **Robin Sequence** | 11/29 (38 %) | 9/13 (69 %) |
|  | **PLSVC** | 5/29 (17 %) | 6/13 (46 %) |
| *Pregnancy/Birth* | IUGR/SGA | 16/26 (62 %) | 10/12 (91 %) |
|  | Preterm birth | 19/26 (73 %) | 11/12 (92 %) |
| *Skeletal* | Growth retardation | 13/22 (59 %) | - |
|  | Scoliosis/vertebral anom. | 8/29 (10 %) | 3/13 (23 %) |
|  | Hand or foot malform. | 3/29 (10 %) | 3/13 (23 %) |
|  | 2-3 toe syndactyly | 6/29 (21 %) | 3/13 (23 %) |
| *Pulmonary* | Respiratory issues | 16/29 (55 %) | 11/12 (92 %) |
| *Heart<sup>A</sup>* | Other | 9/29 (31 %) | 3/13 (23 %) |
| *Urogenital* | Renal anomaly | 6/29 (21 %) | 6/13 (46 %) |
|  | Cryptorchidism | 11/29 (38 %) | 2/13 (15 %) |
| *Vision/Hearing* | ONH/CVI | 14/29 (48 %) | 4/8 (50 %) |
|  | Other ophth. Features | 19/29 (66 %) | - |
|  | Hearing impairment | 9/29 (31 %) | 2/8 (25 %) |
| *Neurodevelopment* | Moderate to severe ID | 15/15 (100 %) | - |
|  | Severe speech delay | 21/21 (100 %) | - |
|  | Severe motor delay | 18/19 (95 %) | - |
|  | Hypotonia | 18/29 (62 %) | 6/10 (60%) |
|  | Epilepsy | 6/29 (21 %) | - |
| *Premature death* | Early childhood | - | 13/13 100 % |
|  | After 3 years | 3/29 (10 %) |  |
|  |  | **TARPS** |  |
| *Gastrointestinal* | Failure to thrive | 26/38 (68 %) |  |

|  |  |  |
| --- | --- | --- |
|  | Tube feeding | 20/38 (53 %) |
|  | VDR/intestinal malrotation | 4/42 (10 %) |
|  | Imperforate anus | 2/42 (5 %) |
| *Brain* | Cerebellar anomaly | 15 /31 (48 %) |
|  | Ventriculomegaly or dilated IV. ventr. | 13/31 (42 %) |
|  | Abnormal gyri | 8/31 (26 %) |
|  | Corpus callosum hypoplasia | 7/31 (23 %) |
|  | White matter anom. | 5/31 (16 %) |
|  | Signs of previous bleeding/sinus thrombosis | 7/31 (23 %) |
|  | Other | 11/31 (35 %) |
| *Characteristic facial dysmorphism* |  | Low set ears, high arched broad eyebrows, up slant often short pf, wide mouth with downturned corners, Robin Sequence, or microretrognathia. |

TARPS (TARP Syndrome) is when relevant subdivided into cTARP (classical TARP syndrome) (13 males), constituting the most severe end of the spectrum and TARPL (TARP-like) (29 males). Features from the TARP acronym are indicated in **bold**. ASD (atrial septal defect), PLSVC (persistent left superior vena cava), IUGR (intrauterine growth retardation), SGA (small for gestational age), ONH (optic nerve hypoplasia), CVI (central visual impairment), VDR (vitelline duct remnant), ID (intellectual disability). Data for cerebral MRI was based on 31 individuals with TARPS (7 cTARP, 24 TARPL). <sup>A</sup>Other heart malformations than ASD and PLSVC.

The number of features defined by the TARP acronym (Talipes, Atrial septal defect, Robin sequence, and Persistent vena cava superior) were larger for individuals with cTARP than for individuals with TARPL (Figure 2A+B). Thus, in cTARP, the average number of TARP-features was 2.4, with all having at least one feature, and the majority (85%) had two or more features (two had all four features). For TARPL, the average number of TARP-features was 1.3, and 45% had two or more features, while 17 % had no features as described by the TARP acronym (Figure 2A+B). The number of features from the TARP-acronym thus increase with increasing severity of the phenotype.

**Figure 2.**
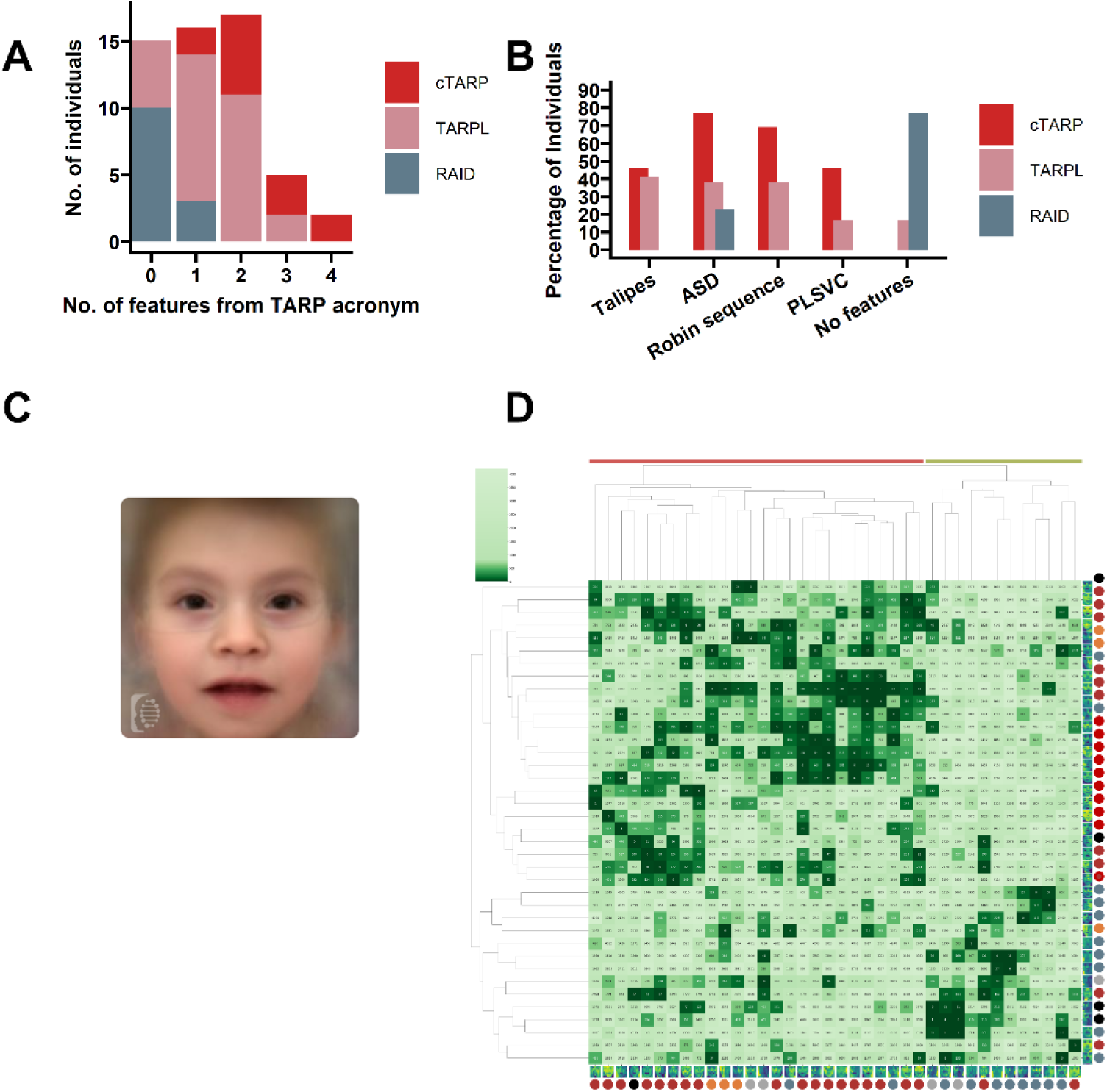
*RBM10* Phenotype Characterization: **A**. The number of features from the TARP acronym (Talipes, Atrial septal defect, Robin sequence, Persistent left superior vena cava) in 55 individuals according to phenotype (13 with cTARP (classical TARP), 29 with TARPL (TARP-like), and 13 with RAID (RBM10 associated intellectual disability)). It can be seen that the number of features is larger in individuals with more severe phenotypes. **B.** Distribution of features from the TARP acronym according to phenotypes. **C.** Composite photo based on 17 images of TARPS (TARP syndrome) individuals. The most frequent dysmorphic features are brachycephaly or other cranial anomaly, low set ears, arched and broad eyebrows, up slanted and often short palpebral fissures, hypertelorism, and wide mouth with downturned corners, and retrognathia. **D.** Clustered heatmap of Ranked matrix based on Face2Gene comparison of images from individuals with *RBM10* variants. Light green indicates low similarity between individuals, dark indicates high similarity. Two major clusters were formed, highlighted with red and green. Images from 8 individuals previously reported in the literature were included in the analysis (15, 16, 18–20, 24, 25, 29). Dark red – TARPS, Orange – Intermediate, Blue – RAID, Black – Unclassifiable phenotype, Grey – Variant of unknown significance.

The frequency of phenotypic features can be seen in Table 1 and Supplementary Table S1. Frequent features across the TARPS were preterm birth, IUGR/low birth weight, and respiratory problems. Failure to thrive was frequent, often with a need for tube feeding. Developmental delay or intellectual disability (ID) were seen in all and were mostly severe. Hypotonia was frequent, epilepsy was seen in some. The majority achieved independent walking late, most after 4 years of age, while some individuals walked only with support or were non-ambulatory. Speech delay was pronounced with most having very poor language with few or no words. Skeletal features were postnatal growth retardation, scoliosis or vertebral anomalies, while malformations of hand or feet were seen in a fraction. Atrial septal defect was the most frequent cardiac malformation (Figure 2B), but other structural anomalies, as ventricular septal defect, were also frequent. Hypertrophic cardiomyopathy was reported in few individuals.

Visual impairment was frequent (usually optic nerve hypoplasia, but also central visual impairment, myopia, nystagmus and strabismus), as was hearing impairment. Genitourinary anomalies were mostly structural renal anomalies and cryptorchidism.

Causes of death in cTARP syndrome (Supplementary Table S2) was mostly respiratory insufficiency. Some had pulmonary hypoplasia secondary to oligohydramnios. Importantly, intestinal obstruction due to vitelline duct remnants, which was fatal in some individuals, is potentially treatable but difficult to diagnose, especially in individuals with severe cognitive impairment.

Dysmorphic features were characteristic and consisted of brachycephaly or other cranial anomaly, low set ears, high arched eyebrows, up slanted palpebral fissures, a wide mouth with downturned corners, and retrognathia or micrognathia. Cleft palate was also frequent with Robin sequence being one of the TARP acronym features. The dysmorphic features were sufficiently characteristic to allow for creation of a facial gestalt for TARPS via Face2Gene (FDNA Inc, USA) (Figure 2C) (40, 41).

Cerebral MRI revealed the most frequent anomalies to be hypoplastic cerebellar vermis, dilation of the 4^th^ ventricle/megacisterna magna, ventriculomegaly, thin or absent corpus callosum, abnormal gyri, and white matter anomalies.

The most frequent prenatal features were increased nuchal translucency, IUGR, cardiac, renal and cerebral anomalies, micrognathia, talipes, and oligohydramnios. Birth was frequently induced due to IUGR and fetal distress.

### RBM10-Associated Intellectual Disability, RAID

Thirteen individuals, 12 from our study (from 9 families), and one previously published individual (29), were categorized as having RAID phenotype (Table 2, Supplementary Table S1). RAID differs from TARPS, in the developmental delay/cognitive abnormality being milder, and absence of multiple organ anomalies. While most individuals with RAID did not have any of the TARP features, three had mild atrial septal defects (Figure 2A+B). Motor development was normal to mild/moderately delayed with all individuals walking before 24 months. Most had speech delay, which was pronounced in some individuals. Cognitive affection was observed in all and ranged from mild to moderate ID. Behavioral anomaly as ADHD, aggressive behavior or autistic features was frequent. Epilepsy was noted in some. Short stature and macrocephaly were present in some. It was not possible to delineate a characteristic facial dysmorphisms. It is, however worth noting that cranial anomaly was frequent, and that some had a small mouth.

**Table 2:** Phenotype of 13 Individuals with RAID.

|  |  | **RAID** |
| --- | --- | --- |
| *TARP Features* | **Talipes** | 0/13 (0%) |
|  | **ASD** | 3/13 (23 %) |
|  | **Robin Sequence** | 0/13 (0 %) |
|  | **PLSVC** | 0/13 (0%) |
| *Neurodevelopment* | Mild/moderate ID | 10/10 (100 %) |
|  | Speech delay | 13/13 (100 %) |
|  | Behavioral anom. | 8/12 (67 %) |
|  | Epilepsy | 3/13 (23 %) |
| *Skeletal* | Short stature<sup>A</sup> | 5/13 (38 %) |
|  | 2-3 toe syndactyly | 4/13 (31 %) |
| *Facial dysmorphism* | Macrocephaly<sup>A</sup> | 6/12 (50 %) |
| Facial dysmorphism is less characteristic |  |  |

Features from the TARP acronym are indicated in **bold**. <sup>A</sup>Macrocephaly and short stature are associated with missense variants in RRM2 only. RAID, (RBM10 associated intellectual disability), ASD (atrial septal defect), ID (intellectual disability).

Five individuals with a phenotype that may be compatible with RAID have variants of uncertain significance and are not included in the summary (P4, P27, P31A, P31B, P31C).

### Intermediate Phenotype

Five individuals, four from our study (from three families) and one from the literature (17), have features from TARPS as well as RAID, and we classify them as having an intermediate phenotype (Supplementary Table S1). They had neurodevelopmental development similar to RAID, but also features as atrial septal defects that had to undergo surgery, optic atrophy, hearing impairment or scoliosis, which are typical for TARPS.

### Facial Gestalt Image Analysis

The phenotype grouping was supported by analysis of facial dysmorphism using the DeepGestalt and GestaltMatcher algorithms in the Face2Gene platform (40) (41) (Figure 2D). We analyzed photos from 29 individuals in our cohort and 9 individuals from the literature (15, 16, 18–20, 24, 25, 29) for similarities in facial anatomical features. Clustering of the ranked matrix revealed two main clusters (Figure 2D). The vast majority of individuals with RAID are placed in a smaller cluster separating from the larger cluster of individuals with TARPS. These results are consistent with our categorization of phenotypes into two groups.

### Genotype-Phenotype Correlation

Fifty-two different variants from 54 families are included in the study (Figure 1A, Table 3 and Supplementary Table S3), 26 variants are reported here for the first time. The RBM10 variant occurred *de novo* in 10/54 of the families and was maternally inherited in the remaining 44 families of which 13/44 were mosaic or had arisen *de novo* in the parent. Missense variants are the most frequent (22/52), followed by frameshift variants (14/52), nonsense variants (8/52), splicing variants (7/52), and large deletions (1/52). Clusters of missense variants are located in the RRM2 domain and in the C_2_H_2_-type ZF domain. A cluster of variants causing a premature termination codon (PTC), are located in the alternatively spliced exon 4. Four missense variants in RRM2 (all substitutions to proline) are observed in individuals with more severe phenotypes (p.L307P (P6) (with cTARP), and p.S315P (17), p.L350P (P14B), and p.H367P (P3) (with an intermediate phenotype)). Two nonsense variants located close to the 5’- or 3’-end (p.R32* (P29), p.Q891* (P21)) are associated with a milder phenotype (intermediate phenotype or RAID respectively). This is most likely due to reinitiation of translation from a downstream in frame start codon (p.R32* (P29), Supplementary Table S3), and creation of a partly functional protein truncated near the C-terminus (p.Q891* (P21)). An overview of the genotype-phenotype correlation can be seen in Table 3.

**Table 3:** Genotype-Phenotype Correlation.

|  | RAID | Inter-<br>mediate | TARPS |  | Unable to clas-<br>sify phenotype<sup>A</sup>/<br>VUS<sup>B</sup> |
| --- | --- | --- | --- | --- | --- |
|  |  |  | TARPL | cTARP |  |
| **RRM2 missense variants** | p.R332C<br>**p.R332H**<br>p.R343C<br>p.L350P | p.H367P<br>p.S315P<br>(17)<br>p.L350P |  | p.L307P | p.P322L (16)<sup>A</sup> |
| **CH2-ZF2 mis-**<br>**sense variants** |  |  | p.C761Y<br>p.R766C<br>**p.R766H**<br>p.H784R |  | p.L774P (34)<sup>A</sup><br>p.R776W(36)<sup>A</sup> |
| **Other missense variants** | p.C222R<br>p.R817C<br>p.W658C<br>(29) |  | p.D755N (26)<br>p.R387K (39) |  | p.R9H<sup>B</sup><br>p.R30C<sup>B</sup><br>p.G625R<sup>B</sup> |
| **Splice variants** | c.432+5G>T |  | c.17+1G>A (30)<br>c.724+2T>C (20)<br>c.901+1G>A,<br>c.1249-1G>A<br>c.1693+4A>C<br>(15)<br>c.2100+1G>A<br>(32) |  | c.724+2C>T (38)<sup>A</sup> |
| **PTCs in exon 4** |  | p.Q138* | p.S73Wfs*60 (15)<br>p.P74Rfs*60 |  |  |
|  |  |  | p.G92Afs*39 (24)<br>**p.R98\***<br>p.E119Mfs*14<br>(33)<br>p.A127Pfs*7 |  |  |
| **PTCs outside**<br>**exon 4** | p.Q891* | p.R32* | p.K54Sfs*80 (19)<br>p.A319Pfs*16<br>p.S602Pfs*25<br>(25)<br>p.S738Rfs*63 | p.Q150* (20)<br>p.I372Afs*110<br>(28)<br>p.W412* (14)<br>p.E451Vfs*66<br>(21)<br>p.H626Lfs*78<br>(27)<br>**p.P632Tfs\*41**<br>(14) (the original<br>family of<br>(18)),(23)<br>p.R680* (31)<br>p.R726* (20) | p.Asp847Leufs*12<br>(37)<sup>A</sup> |
| **Large deletions** |  |  | p.651_889del<br>(10) |  |  |

RBM10 variants from this study and published variants are categorized according to phenotype.
Variants seen in two or more unrelated individuals are in **bold**. Variants, which we were not able to classify<sup>(A)</sup> and VUS<sup>(B)</sup> (variants of unknown significance) are indicated in grey in the last column.
RAID (RBM10 associated intellectual disability), TARPS (TARP Syndrome), TARPL (TARP like), cTARP (classical TARP syndrome), PTC (premature termination codon). Protein reference: NP\_005667.2.

**Table 4:**
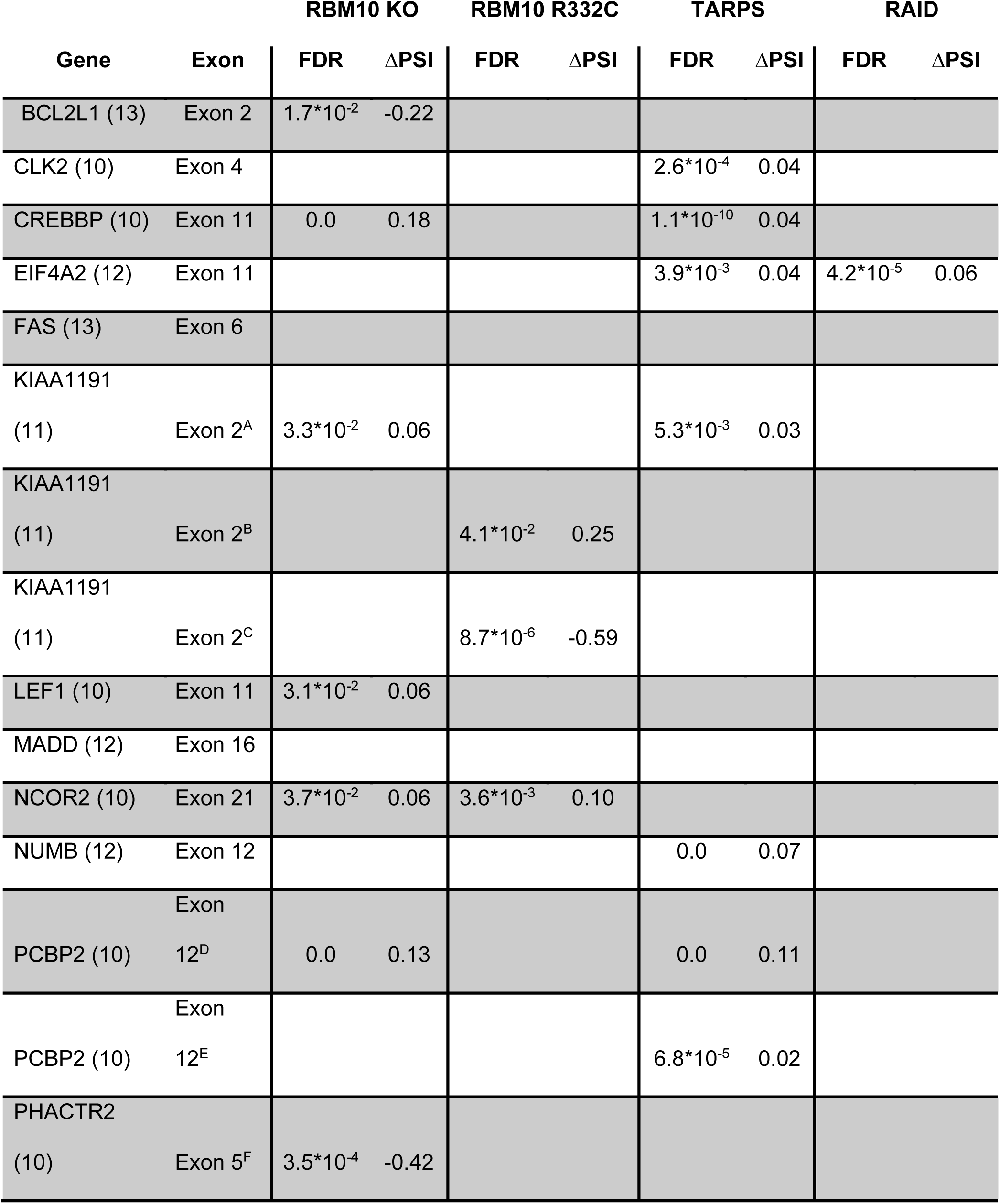
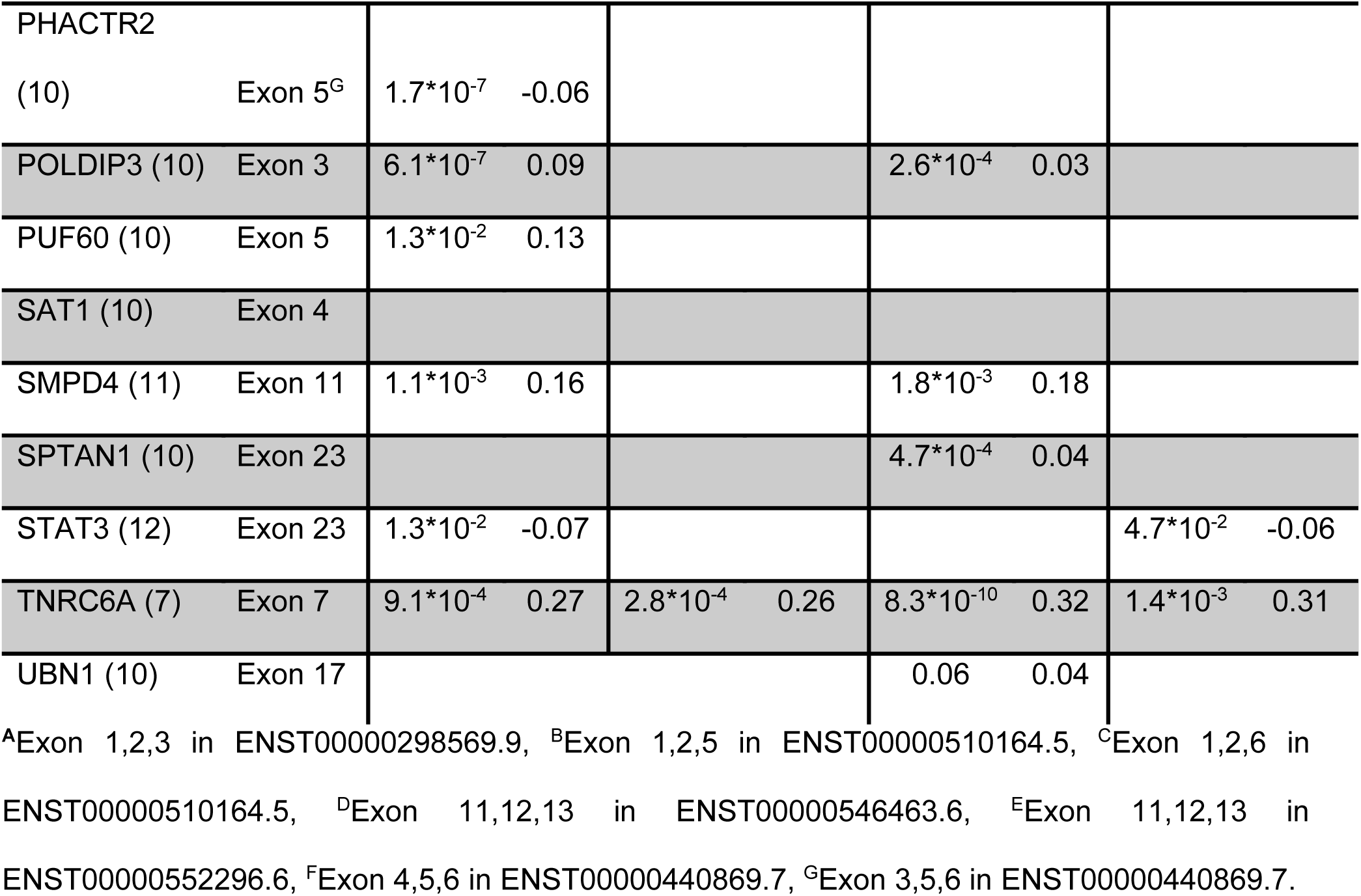
Splicing of Known RBM10 Targets.

It is evident that a genotype-phenotype correlation exists. Among 10 families with cTARP 8 have LOF variants located outside exon 4, and only one has a missense variant. In 26 families with TARPL there are 23 different variants (in one family the RBM10 variant was not indicated): ten PTC variants (six PTC variants cluster within exon 4), six splice site variants, one large deletion, and six missense variants (four cluster in the C_2_H_2_-type ZF domain). In five families with intermediate phenotype there are five different variants: three missense (substitution to proline in RRM2) and two atypical non-sense variants (Supplementary Table S3). In 10 families with RAID there are nine different variants: seven missense variants (four cluster in the RRM2 domain), one splice site variant, and one non-sense variant located close to the C-terminus.

### Identifying Functional Consequences of Variants in RBM10 Individuals

To characterize the functional consequences of the *RBM10* variants, we performed RNA-seq on blood samples from 12 individuals from 12 families (Supplementary Table S3). Eight samples from male relatives were used as healthy controls. Individuals were grouped into the two main phenotype groups. Gene expression analysis identified 243 genes (Supplementary Table S4) with significant (padj-value <0.05) changes in gene expression in TARPS individuals (n=8), 135 genes were upregulated and 108 were downregulated. In the RAID individuals (n=4) only 27 genes (Supplementary Table S5) had a significant change in gene expression, 11 genes were upregulated, and 16 were downregulated. Four of these overlapped with significant altered genes in TARPS and were among the most up- or downregulates genes. The low number of significant genes in RAID indicates that the effects of the *RBM10* variants present in the RAID group are less severe. This was also evident from the splicing analysis, where 3545 significant (FDR <0.05) splicing events were identified in the TARPS group and only 1901 events in the RAID group (Figure 3A+B). Alternative splicing of CE was the most frequent type of alternative splicing observed. In TARPS individuals, 563 CE were significantly more skipped, and 1433 were more included (Figure 3A, Supplementary Table S6). In RAID individuals, 433 CE were significantly more skipped, and 612 were more included (Figure 3B, Supplementary Table S7). Our analysis shows that the *RBM10* variants more frequently increase exon inclusion, which is consistent with the known function of RBM10 as a negative regulator of CE (10, 12), suggesting that the variants mainly cause loss of RBM10 function.

**Figure 3.**
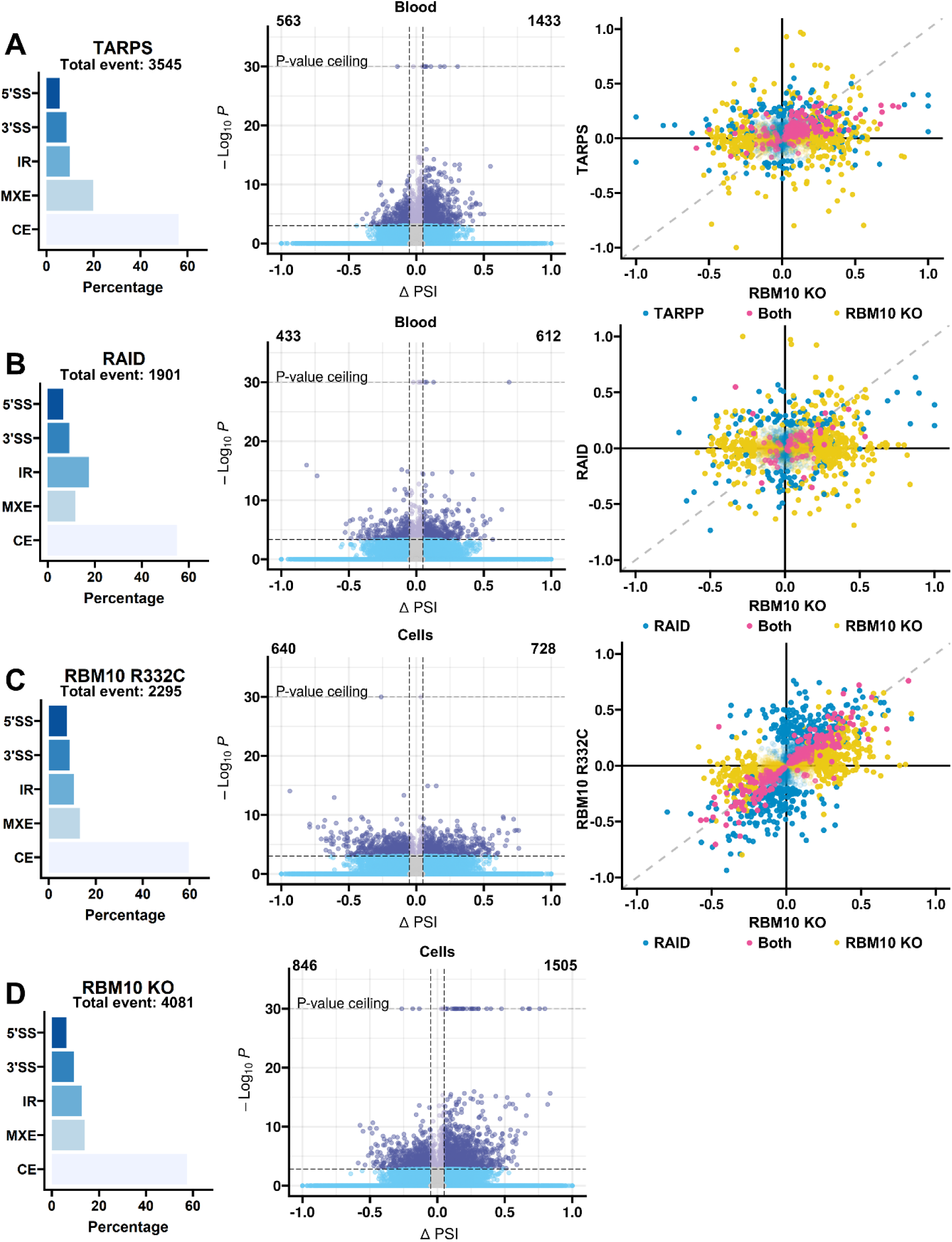
RBM10 Splicing Regulation Analysis: RNA-sequencing splicing analysis on RNA from blood or RBM10 CRISPR edited cells compared to control. Left – Distribution of significant (FDR <0.05) changed alternative splicing events. 3’SS (Alternative 3’ splice site), 5’SS (Alternative 5’ splice site), IR (Intron retention), MXE (Mutually exclusive exon), CE (Cassette exon). Middle – Volcano plot visualizing Δ percent spliced in (PSI) for significant CE events. Right – Pairwise comparison of ΔPSI for significant CE in the different data sets compared to RBM10 KO. Blue dots - significant in the analyzed group, pink dots – significant in both groups, yellow dots – significant in RBM10 KO. Events significant in one data set with a ΔPSI <0.2 are visualized as transparent dots. **A.** RNA from blood from TARPS (TARP syndrome) (n=8) compared to healthy controls (n=8). **B.** RNA from blood from RAID (RNA associated intellectual disability) (n=4) compared to healthy controls (n=8). **C.** RNA from CRISPR R332C cells (n=3) compared to WT cells (n=4). **D.** RNA from CRISPR KO cells (n=6) compared to WT cells (n=4).

To identify targets and splicing events associated with different degrees of RBM10 LOF, we established three different RBM10 CRISPR edited cell lines: two KO (KO1 and KO5) cell lines (to mimic TARPS), and one cell line harboring a p.R332C missense variant in the RRM2 domain (R322C cell line), associated with a RAID phenotype in two individuals (P1A and P1B) (Supplementary Figure S1). Employing RNA-seq analysis, we compared gene expression and splicing patterns to those of WT RPE-1 cells. In RBM10 KO cells we observed 442 genes with significant changes in gene expression (244 genes were upregulated and 198 were downregulated (Supplementary Table S8)). Further, 4081 significantly altered splicing events were identified (Figure 3D) of which 2352 were CE events, with 1506 exons being more included and 846 exons more skipped. This further supports the notion that loss of RBM10 function mainly increases inclusion of CE (Figure 3D, Supplementary Table S9, ratio of skipping/inclusion: 0.34/0.64). Pairwise comparison of significantly changed CE between KO cells and samples from TARPS individuals identified 266 overlapping events (Supplementary Table S10). The majority of these events were increased CE inclusion in both data sets (Figure 3A).

In the cell line harboring the RAID associated p.R322C variant, only 91 genes showed significant altered gene expression (44 upregulated and 47 downregulated, Supplementary Table S11). Splicing analysis identified 2296 significantly altered splicing events of which 1368 were altered CE events, 640 were more skipped and 728 were more included (ratio of skipping/inclusion: 0.47/0.53, Supplementary Table S12). Pairwise comparison between the KO cell lines and samples from RAID individuals identified only 69 overlapping events (Figure 3B, Supplementary Table S13).

Taken together our analysis shows that the *RBM10* variants in the TARPS group generally result in more splicing alteration events than the variants in the RAID group (ratio of skipping/inclusion: 0.28/0.72 vs 0.41/0.59). The overlap in CE splicing events between RAID and TARPS individuals and the KO cell lines, demonstrates that both phenotype groups have loss of RBM10 protein function, and that the loss of RBM10 activity is more severe in TARPS individuals than in RAID individuals, consistent with the more severe phenotype observed in TARPS individuals.

### Altered Protein Activity Drives LOF

Our RNA-seq analysis demonstrated that disease severity is correlated with the degree of RBM10 LOF in the two phenotype groups. To investigate the molecular consequences of the missense variants underlying the different phenotypes, we first investigated the levels of full-length RBM10 protein in two available patient fibroblast cell lines, one from an individual with cTARP harboring an RRM2 variant (p.L307P, (P6)) and another one from an individual with TARPL harboring a C_2_H_2_-type ZF domain variant (p.R766H, (P8)).

Fibroblasts with the p.L307P variant displayed severely reduced levels of RBM10 protein, whereas RBM10 protein amounts in fibroblasts from the individual with the p.R766H variant were comparable to control WT RBM10 fibroblasts (Figure 4A). *RBM10* mRNA expression levels were unaltered in both cell lines (Supplementary Figure S2).

**Figure 4.**
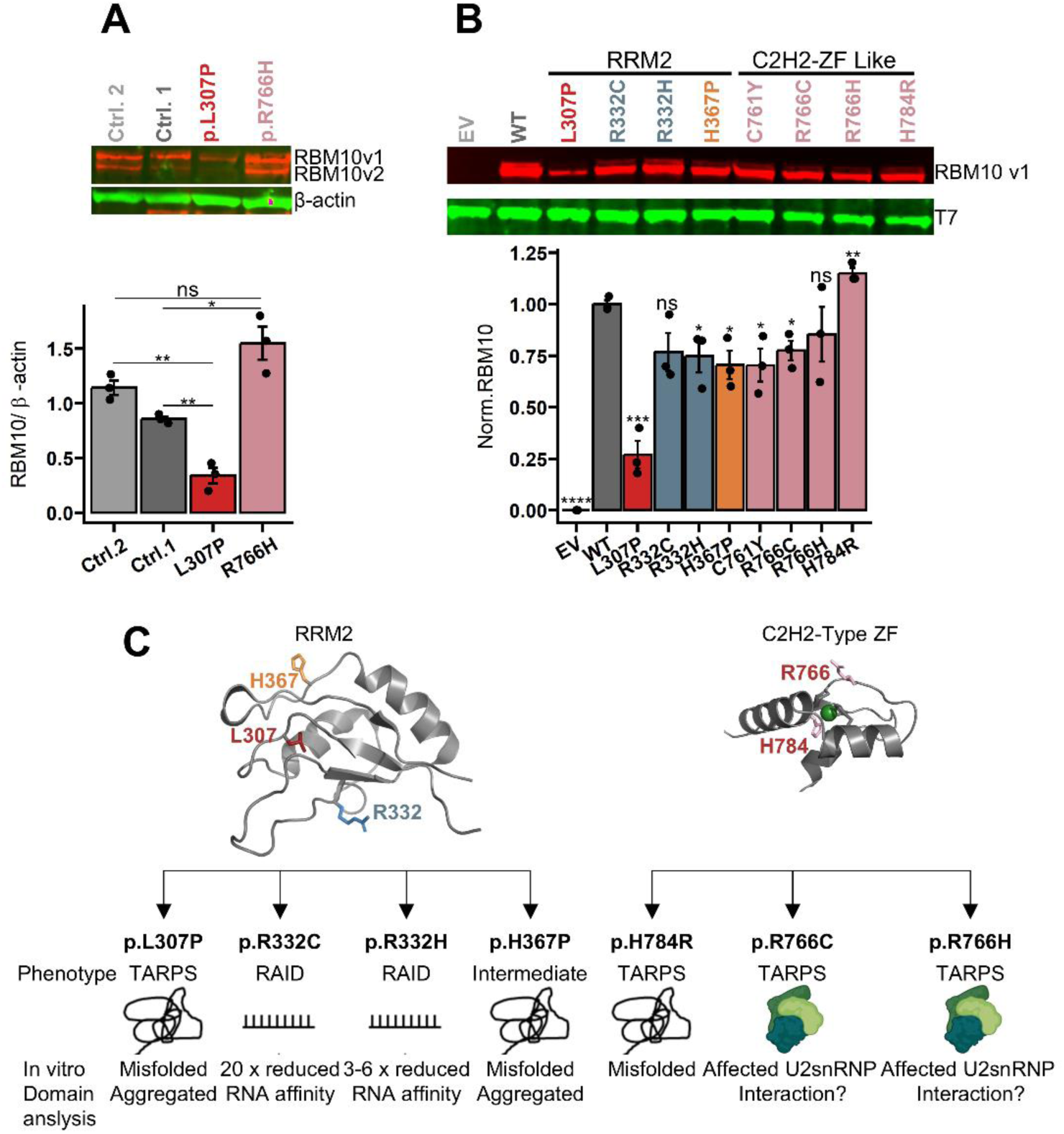
Protein and Domain Investigation in Mutant RBM10. **A.** Western blot and quantification (n=3) representing full-length RBM10 protein in RBM10 fibroblasts from an individual with a TARP-Like (TARPL) variant in the C_2_H_2_-type ZF domain (p.R766H, P8), an individual with classical TARP (cTARP) and an RRM2 domain variant (p.L307P, P6) and two control fibroblasts from healthy individuals. Error bars represent calculated SEM (Standard error of mean). **B)** Western blot and quantification of full-length RBM10 protein in CRISPR KO cells transfected with RBM10v1 expression vectors harboring variants from individuals with RBM10 associated syndromes, compared to WT RBM10v1 and empty vector. Four variants from the RRM2 domain (p.L307P, p.R332C, p.R332H, p.H367P) and four variants from the C_2_H_2_-type ZF domain (p.C761Y, p.R766C, p.R766H, p.H784R). Relative amount of RBM10 protein normalized to total protein and T7-vector used as transfection efficiency control (n=3). Error bars represent calculated SEM. **C)** Summery of mutational consequences based on *in vitro* domain construct analysis of seven selected *RBM10* variants in RRM2 or C_2_H_2_-Type ZF domain constructs. Listed below the variant is the correlated phenotype and observed consequence of the variant in domain construct. RAID (RBM10 associated intellectual disability), TARPS (TARP syndrome). *p<0.05, **p<0.01, ***p<0.001, ****p<0.0001 by two-sample, two-tailed *Student’s t-test* assuming equal variances.

These results were mirrored by RBM10 over-expression analysis in RBM10 KO cells where we analyzed four C_2_H_2_-type ZF domain variants (p.C761Y, p.R766C, p.R766H, and p.H784P) identified in individuals with TARPL, two RRM2 variants from individuals with RAID (p.R332C and p.R332H), one RRM2 variant from an individual with an intermediate phenotype (p.H367P) as well as the severe p.L307P RRM2 variant. Consistent with the results from patient fibroblast we observed comparable amounts of RBM10 protein from the p.R766H variant and severely decreased protein amounts from the p.L307P variant (Figure 4B). Moreover, protein amounts from the three other C_2_H_2_-type ZF domain variants and the other three RRM2 domain variants were slightly reduced compared to WT (Figure 4B). We therefore hypothesized that these variants affect the structure or molecular interactions of RBM10, leading to a less functional protein and/or a protein with altered functions. To functionally characterize the mutant proteins in more detail, we analyzed the structure, stability of Zn^2+^ coordination, and RNA-binding affinity employing recombinant proteins comprising the RRM2 and the C_2_H_2_-type ZF domains of RBM10 harboring selected patient variants (Figure 4C, Supplementary Table S3).

We used nuclear magnetic resonance spectrometry (NMR) analysis to assess the integrity of the three-dimensional structure to characterize the effect of RBM10 variants. Comparison of NMR spectra of the three C_2_H_2_-type ZF mutant versions with the corresponding WT protein (residues 726-820) showed that only the p.H784R variant directly affects the structure of the domain, while the spectra of the other two variants (p.R766C and p.R766H) are comparable to the wild type (Supplementary Figure S3A). The p.H784 mutant protein is involved in the Zn^2+^ coordination, and replacement of histidine with arginine leads to misfolding of the zinc finger and, therefore, a non-functional C_2_H_2_-type ZF domain. According to the predicted structure, p.R766 lays on the surface of the domain but in the vicinity of the Zn^2+^ coordination sphere (Figure 4C). We hypothesized that substitution of p.R766 to histidine or cysteine would alter the Zn^2+^ coordination stability as has been shown for other zinc fingers with consecutive cysteine residues (42). Zn^2+^-Cd^2+^ exchange experiments indicated only a slightly reduced stability of the p.R766 mutants, much less than observed for the p.H784R mutant (Supplementary Figure S3B). This suggests that the variants affecting p.R766 do not affect the folding but rather the interactions with binding partners of the domain. In fact, the C_2_H_2_-type ZF domain of RBM10 has recently been demonstrated to be essential for the interaction with the U2 snRNP particle in chromatin-associated splicing complexes (9). These mechanisms are consistent with the LOF effects identified in the RNA-seq analysis and suggest that the loss of protein function resulting either from misfolding (p.H784R) or altered molecular interactions (p.R766C and p.R766H).

To investigate in more detail the effects of four variants (p.L307P, p.R332C, p.R332H, p.H367P) located in the RRM2 domain, we expressed, purified, and compared their structure and RNA binding with the WT version (Figure 4C). Recombinant RRM2 domain (residues 300-384) harboring p.L307P and p.H367P variants formed insoluble aggregates, which could not be refolded. This indicates that these variants cause severe misfolding of the domain, consistent with the rather dramatic amino acid change to proline. Interestingly, only the p.L307P showed a corresponding drastic reduction of protein levels in fibroblast and cell lines, indicating that for this variant severe RRM2 domain misfolding causes destabilization of the entire protein, whereas p.H367P causes local misfolding of the domain without affecting the stability of the entire RBM10 protein (similar to local misfolding of the C_2_H_2_-type ZF domain in p.H784R). The p.R332C and p.R332H variants were soluble and showed NMR spectra comparable to wild type with only minor differences for signals assigned to residues surrounding the mutated position 332 (Supplementary Figure S4A). The RRM2 of RBM10 and its homolog RBM5 are known to interact with single-stranded RNAs, where residue R332 may be involved in the establishment of electrostatic interactions with the negatively charged nucleic acid (7, 43, 44). We used NMR and isothermal titration colorimetry (ITC) to test the interaction of RRM2 variants with RNA sequence motifs derived from the *NUMB* sequence, a known target of RBM10 (12)(Supplementary Figure S4B+C). The p.R332H variant, which maintains a positive charge, showed a 3-6 fold decrease in RNA binding affinity compared to wild type. In contrast, the p.R332C variant, which abolishes the positive charge, showed a more than 20-fold reduced affinity to the same target. Thus, the phenotypes observed in individuals with variants in this codon are most likely caused by the altered RNA binding, likely resulting in a modified binding pattern.

Taken together these results indicate that the variants can cause different degrees of LOF by various mechanisms. The p.L307P variant results in nearly complete LOF due to severely reduced protein levels, causing cTARP, whereas missense variants in the C_2_H_2_-type ZF domain presumably result in a reduced RBM10 function in U2 snRNP recruitment, all associated with TARPL. Finally, the variants that impair or modify the RNA binding ability of RRM2 cause RAID/intermediate phenotypes (Figure 4D).

### Different Degrees of LOF are Reflected in the Splicing Efficiency of TNRC6A exon 7

RBM10s role as a splicing regulator and its regulated targets has been studied extensively. We first analyzed the splicing patterns of 43 validated RBM10 target genes in RBM10 KO and RBM10 R332C cell lines, and in samples from TARPS and RAID individuals (Table 3, full list of targets in Supplementary Table S14 (7, 10–13, 45)). Thirteen known RBM10 target splicing events were identified as significantly altered in RBM10 KO cells and 10 of these followed the pattern previously observed from RBM10 knock down (KD) (7, 10–13). Eleven events were identified as significantly changed in TARPS individuals, all following the pattern observed for RBM10 KD (7, 10–12). Only three known RBM10 target events were identified as significantly altered in the RAID group, although many other known targets followed the pattern of RBM10 KD, but only with small and not statistically significant changes (Table S15). This further supports that the loss of RBM10 protein function is less pronounced in the RAID individuals than in individuals with TARPS.

The most pronounced alteration in a known RBM10-regulated splicing event was observed for *TNRC6A* exon 7 (ENST00000395799.8) in all investigated data sets (Figure 5A, Supplementary Figure S5). *TNRC6A* exon 7 is regulated by RBM10 and a PAR-CLIP motif matching the RRM1-RanBP2-ZF binding site has been identified in exon 7 (7). Moreover, a missense variant in *RBM10* RRM2 has previously been shown to affect splicing of this exon (17).

**Figure 5.**
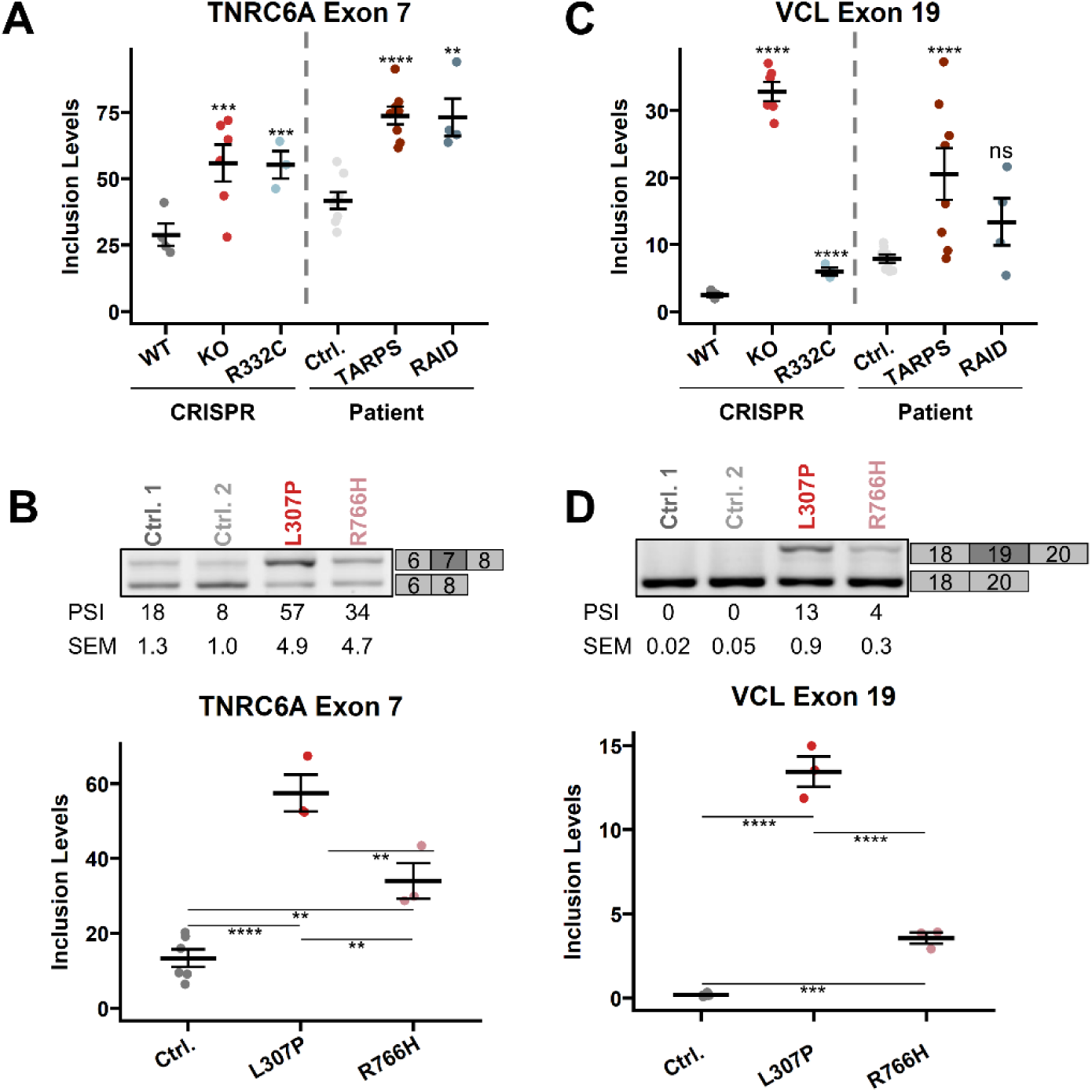
Demonstrating Gradual Loss of RBM10 Protein Function by Splicing Analysis: **A.** *TNRC6A* exon 7 (ENST00000395799.8) inclusion levels in RBM10 KO cells (ΔPSI=0.27, n=6) and RBM10 R332C cells (ΔPSI=0.26, n=3) compared to WT cells (n=4) and in TARPS (TARP syndrome) (ΔPSI=0.32, n=8) and RAID (RBM10 associated intellectual disability) (ΔPSI=0.31, n=4) individuals compared to healthy control n=8. **B.** *TNRC6A* exon 7 splicing in RBM10 fibroblast from a TARPL (TARP-like) individual with a C_2_H_2_-type ZF domain variant (p.R766H, P8), a cTARP variant in the RRM2 domain (p.L307P, P6), and two control fibroblast from healthy individuals (FB01 and FB07). Error bar represents calculated SEM (standard error of mean) (n=3). **C.** *VCL* exon 19 (ENST00000211998.10) inclusion levels in RBM10 KO cells (ΔPSI=0.30, n=6) and RBM10 R332C (ΔPSI=0.03, n=3) cells compared to WT cells (n=4), and in TARPS (ΔPSI=0.13, n=8) and RAID individuals (n=4) compared to healthy control (ΔPSI=0.06, n=8). The change in inclusion level between RAID and control is non-significant (ns). **D.** *VCL* exon 19 splicing in RBM10 fibroblast from a TARPL individual with a C_2_H_2_-type ZF domain variant (p.R766H, P8), a cTARP variant in the RRM2 domain (p.L307P, P6), and two control fibroblast from healthy individuals (FB01 and FB07). Error bar represents calculated SEM (n=3). p-values calculated by two-sample, two-tailed *Student’s t-test* assuming equal variances.*p<0.05, **p<0.01, ***p<0.001, ****p<0.0001

In fibroblasts, *TNRC6A* exon 7 was significantly more included in samples from the individual with the severe p.L307P variant (P6 with cTARP), that causes almost complete loss of RBM10 protein, than in fibroblast from the individual with the milder p.R766H (P8 with TARPL) and in both TNRC6A exon 7 was significantly more included than in controls. These results further demonstrate the correlation between the degrees of LOF and the severity of the clinical phenotype (Figure 5B, Supplementary Figure S5).

### RBM10 Regulated Splicing of Vinculin Exon 19

Vinculin (VCL) is a cytoskeletal scaffolding protein involved in cellular adhesion. *VCL* exon 19 is alternatively spliced and transcripts lacking exon 19 encode the ubiquitously expressed VCL protein, whereas transcripts with exon 19 inclusion encode meta-VCL, which is mainly expressed in smooth-, skeletal-, and cardiac muscles (46). VCL exon 19 was recently identified to be regulated by direct binding of RBM10 (47). Our RNA-seq analysis showed altered splicing of *Vinculin (VCL)* exon 19 (ENST00000211998.10). *VCL* exon 19 was significantly more included in RBM10 KO cells and in TARPS individuals (Figure 5C). This was also observed, to a lesser extent, in the RAID individuals, but did not reach statistical significance (Figure 5C), Supplementary Figure S6). The observed increase in the levels of exon 19 inclusion in the TARPS individuals corresponded to the different degree of loss of RBM10 protein function by their variants (Supplementary Figure S6).

This pattern was also observed in the patient derived fibroblast (Figure 5D), where significantly more exon 19 inclusion was observed in the cTARP (p.L307P, (P6)) fibroblasts with almost complete loss of RBM10 protein compared to the TARPL individual (p.R766H, (P8)) with only partial loss of protein function.

### Consequences of Only Expressing the Exon 4 Skipped RBM10v2 Isoform

A group of individuals with TARPS have variants introducing PTCs within the alternatively spliced exon 4. These individuals all have a TARPL phenotype and not cTARP, despite having PTCs. PTC variants within exon 4 cause the full length RBM10v1 protein isoform to be severely truncated and likely non-functional, or - more likely - the mRNA to be degraded by the nonsense mediated mRNA decay (NMD) pathway (24). We therefore assume that individuals with PTC variants in exon 4, only produce the short RBM10v2 protein isoform, which lacks exon 4. Possible functional differences between the two RBM10 protein isoforms are unknown, but the 16 amino acids encoded by the 3’end of exon 4 correspond to the first part of RRM1 including the RNP2 motif, which is important for classical RNA binding by RRM domains and RRM1 RNA binding (48). We observed that *RBM10* mRNA expression was reduced in individuals with PTC variants in exon 4 compared to healthy controls and the other individuals with *RBM10* variants, likely because the PTCs trigger NMD of the exon 4 containing transcripts (Figure 6A, Supplementary Figure S7(reads from RNA-seq)). The decrease in total amounts of RBM10 can potentially explain the TARPL phenotype, but it could also be caused from missing crucial splicing regulatory functions unique to the RBM10v1 isoform. Interestingly, an individual with a variation in the 5’splice site of exon 4 (c.432+5G>T, (P30)), which results in complete skipping of exon 4 (Figure 6B) was classified to have RAID. This individual exclusively expresses the short RBM10v2 isoform. Consistent with the fact that the c.432+5G>T variant does not result in introduction of PTCs, the expression level of total *RBM10* mRNA was not reduced in this individual. The transcript levels of *RBM10v2* mRNA are equivalent to the sum of v1 and v2 mRNAs in normal individuals because the c.432+5G>T variation shifts splicing to exclusively producing the *RBM10v2* isoform (Figure 6A+C). Splicing of *TNRC6A* exon 7 and *VCL* exon 19 in this individual was unchanged (Figure 6B), suggesting that the splicing regulatory function of the short RBM10v2 isoform is sufficient to maintain correct splicing of these targets. These findings indicate that the TARPL phenotype observed in individuals with PTC variants in exon 4 mainly arise from reduced total amounts of RBM10 protein. However, expression of normal amounts of the short RBM10v2 isoform is insufficient for normal development, as the individual exclusively expressing the short RBM10v2 isoform displays a RAID phenotype (Figure 6C).

**Figure 6.**
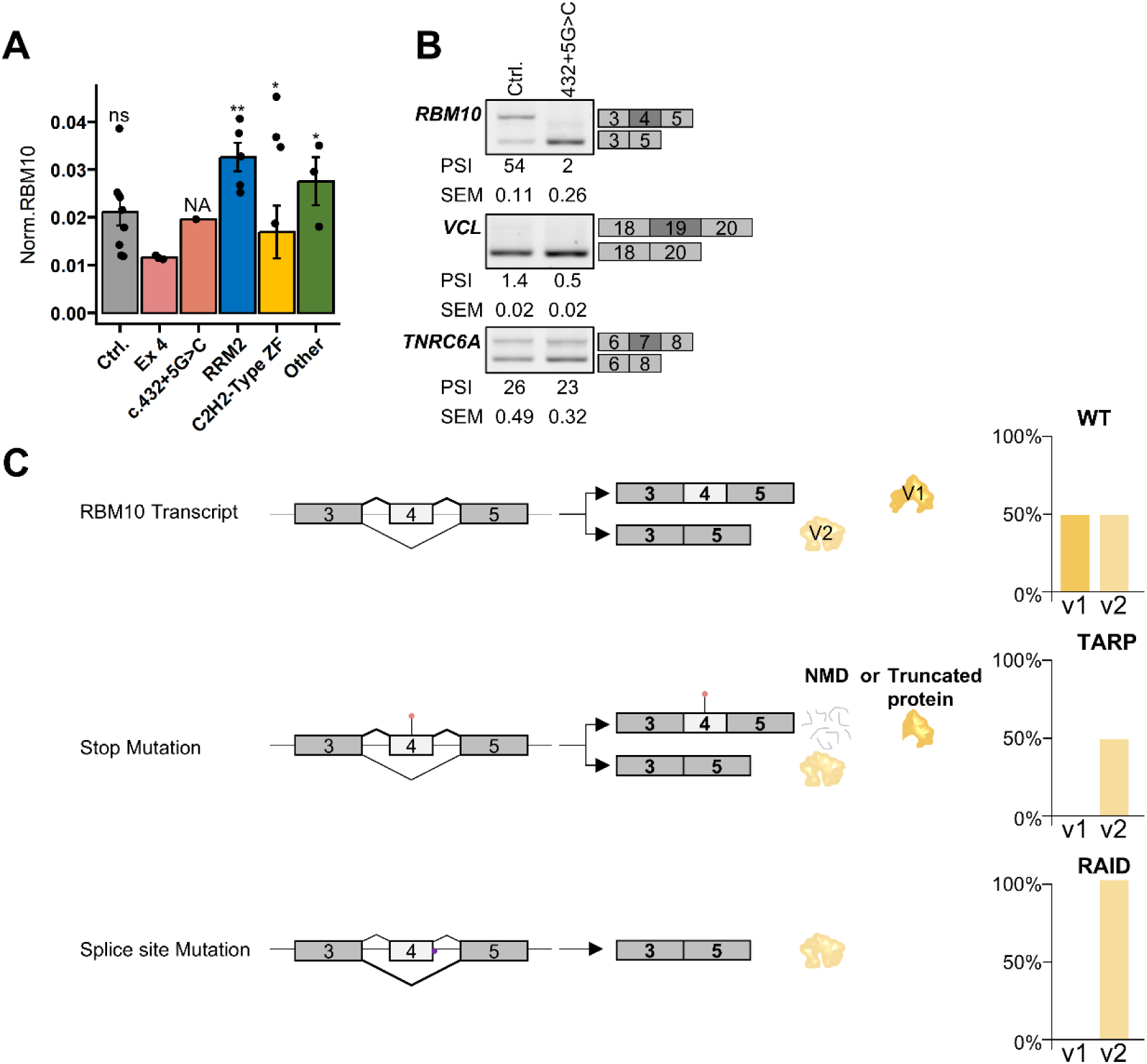
RBM10 Protein Isoform v2 4: **A.** *RBM10* mRNA expression from RNA from blood of individuals with *RBM10* variants, investigated with RT-qPCR normalized to RPL13A and TBP. Ctrl. n=9, Ex 4 n=3, c.432+5G>C n=1, RRM2 n=5, C_2_H_2_-Type ZF n=4, other n=3. *p<0.05, **p<0.01, by two-sample, two-tailed *Student’s t-test* assuming equal variances. **B.** Splicing regulation investigated in an individual with a 5’splice site variant of exon 4, thus only expressing the short RBM10v2 isoform. Splicing in *RBM10* exon 4, *VCL* exon 19, and *TNRC6A* exon 7. PSI (Percent spliced in) and SEM (standard error of mean) calculated from samples of RNA from two blood samples drawn at the same time n=2. **C)** Schematic illustration of the consequences of expressing the different RBM10 protein isoforms. Top – WT *RBM10* resulting in both RBM10v1 and RBM10v2 protein isoforms. Middle – Individuals with STOP variants within exon 4, resulting in *RBM10v1* transcripts to be degraded by the nonsense mediated mRNA decay) (NMD) pathway or expression of truncated protein. This results in an overall decrease in RBM10 protein expression and is associated with TARPL (TARP-like). Bottom – Individuals with exon 4 splice site variants, resulting in full skipping of exon 4 and exclusively expression of the shorter RBM10v2 isoform. Total RBM10 protein expression is unaltered. This is associated with the milder phenotype RAID (RBM10 associated intellectual disability).

### Functional Testing of LOF and GOF Effects

We have demonstrated a clear correlation between varying degrees of LOF on different splicing targets and phenotype severity. We hypothesize that the effects of the sequence variants are not only LOF, but that alterations in the RNA recognition domains could also affect target recognition and have GOF effects, as previously described for other splicing regulatory RBPs (49, 50). To investigate LOF and GOF effects we used splicing reporter mini-genes, co-transfected with RBM10 expression vectors in RBM10 KO cells.

RBM10 can repress splicing when bound close to the 3’ss and may interact with SRSF2 (5). We decided to use the c.362C>T *ACADM* mini-gene as a splicing reporter assay. Here exon 5 inclusion is suppressed due to a weak 3’ss (51, 52). Consistent with RBM10s role as a splicing repressor that functions on weak 3’ss, RBM10 WT, all variant RBM10 proteins, except p.H367P and p.L307P, completely inhibited *ACADM* exon 5 inclusion (Figure 7A). This suggests that on this target all the other variant RBM10 proteins have maintained the ability to mediate strong splicing repression. Interestingly, the p.H367P and the p.L307P proteins conversely resulted in more exon inclusion than observed in the RBM10 KO samples. This suggests that these mutant RBM10 proteins have not simply lost their ability to repress splicing, but instead they are able to stimulate splicing of this target exon. This could be due to altered RNA binding affinity causing a new direct GOF towards this target and likely other similar exons or it could be mediated through an altered interaction with SRSF2, which has also been demonstrated to repress *ACADM* exon 5 inclusion (51, 52).

**Figure 7.**
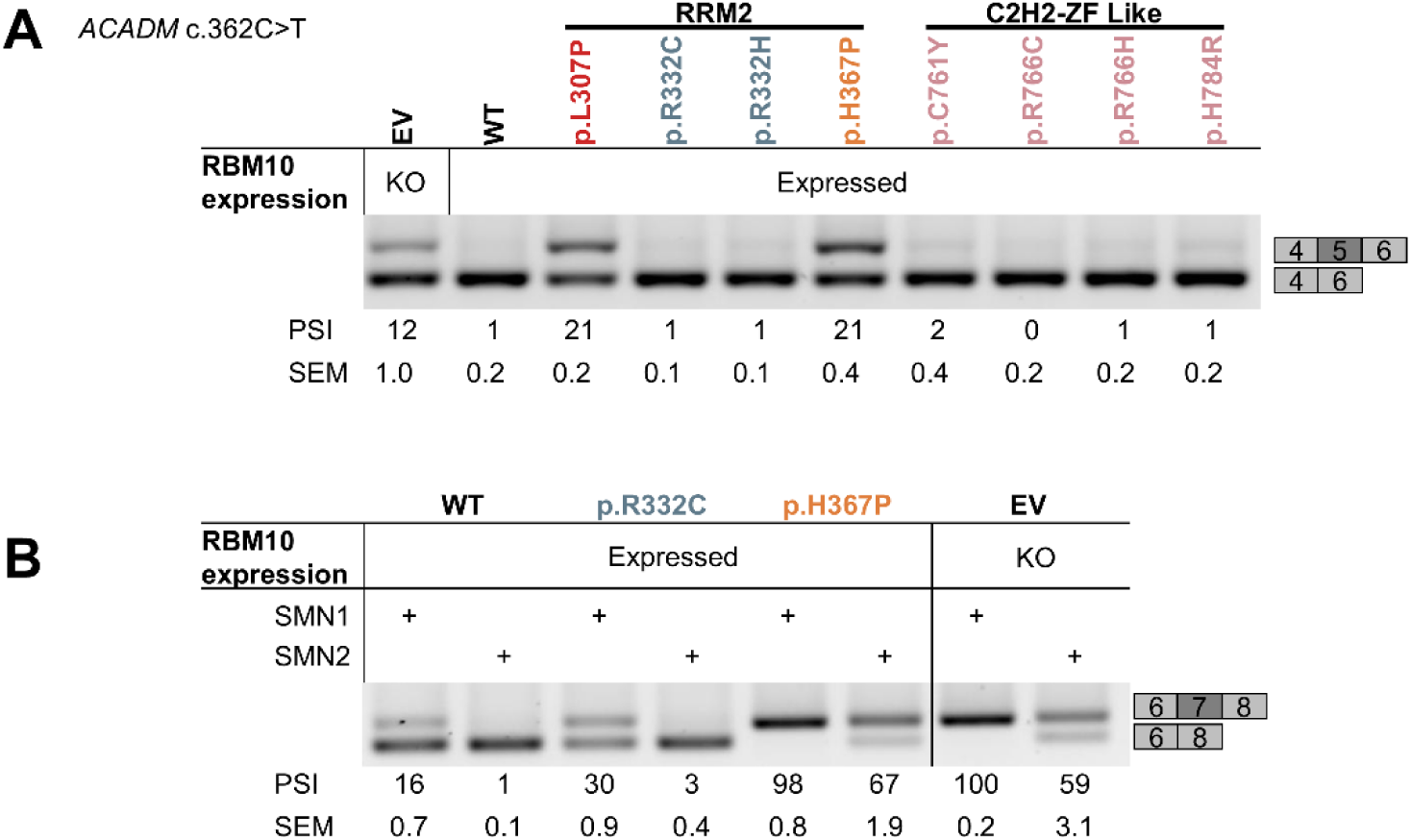
Functional Testing using Splicing Reporter Mini-Genes: **A.** Splicing regulation of *ACADM* exon 5 in a splicing reporter mini-gene with an ESE disrupting variant (c.362C>T), investigated in RBM10 KO cells transfected with RBM10v1 transfection vectors (, p.L307P, p.R332C, p.R332H, p.H367P, p.R766C, p.R766H, and p.H784R). PSI (percent spliced in) and SEM (standard error of mean) (n=3). **B.** *SMN1* and *SMN2* RBM10 regulated splicing of exon 7 investigated with RT-PCR in RBM10 KO cells transfected with RBM10v1 expression vectors, co-transfected with *SMN1/2* splicing reporter mini-genes. Two RRM2 RBM10 variants (p.R332C and p.H367P) with different outcome in phenotype were investigated. PSI and SEM (n=3). Black line marks non-contiguous gel lanes.

To test if this potential GOF effect of the p.H367P protein is general to more targets we also tested its effect on *SMN1* and *SMN2* splicing reporter mini-genes and compared to the p.R332C variant, which had maintained its ability to repress *ACADM* exon 5 (Figure 7B). RBM10 has previously been shown to regulate the splicing of *SMN2* exon 7 in spinal muscular atrophy fibroblasts (11). A single C>T transition in exon 7 of *SMN2* causes almost complete skipping of the exon. The p.R332C variant was identified in individuals with RAID and was shown to affect the RNA binding affinity, whereas the p.H367P variant was identified in an individual with an intermediate phenotype and was shown to cause local misfolding of the RRM2 domain. We show that in cells expressing the p.H367P mutant protein, the splicing of exon 7 in *SMN1* and *SMN2*, was comparable to that observed in the RBM10 KO samples, demonstrating that on this target the p.H367P mutant RBM10 protein has completely lost its ability to inhibit splicing, but without the further GOF splicing stimulatory effect that was observed on *ACADM* exon 5. In samples expressing the p.R332C mutant protein we observe less exon 7 skipping, supporting the notion that the p.R332C protein is less functional than the WT protein, but still retains some residual splicing inhibitory activity. These results support the notion that *RBM10* variants identified in individuals with more severe phenotypes have a splicing pattern mimicking RBM10 KO, whereas *RBM10* variants identified in RAID individuals show less pronounced splicing changes.

Taken together these results corroborate the hypothesis that a RBM10 protein with sequence variants in an RNA binding domain may have LOF towards some targets (e.g. *SMN1/SMN2* exon 7) and GOF towards other targets (e.g. *ACADM* exon 5).

## Discussion

We have characterized 37 not previously published males with *RBM10* variants, and compared them to 34 cases from the literature. Description of phenotypes and functional studies have revealed a clear genotype-phenotype correlation where the phenotypes belong to a “RBM10-phenotypic spectrum”, which can be subdivided into two major groups, TARPS and RAID, based on the severity of symptoms (Supplementary Figure S8).

We suggest that TARPS can be further divided into two subgroups: cTARP defined as those who die soon after birth or in infancy, and TARPL, who constitutes the remaining individuals. Though this subdivision is arbitrary, especially as better treatment options through time has improved survival, this demonstrates that the most severely affected can to some extent be distinguished from the remaining individuals (TARPL) based on the presence of more TARP features and an RBM10 variant causing severe LOF.

The RAID phenotype is milder without involvement of multiple organ systems. For those with RAID caused by missense variants in the RRM2 domain, short stature and macrocephaly are characteristic features.

Some individuals seem to have an intermediate phenotype with features from RAID as well as TARPS. The presence of these individuals supports that the phenotypes are indeed part of one broad common RBM10-phenotypic spectrum.

Phenotype grouping by the next-generation phenotyping tool Face2Gene supported the existence of the two major phenotypes TARPS and RAID. This tool has previously been used to distinguish new syndromes from known syndromes (53) (41). Besides supporting that the spectrum consists of two separate groups, the analysis confirms that the facial dysmorphism is characteristic and recognizable for the TARPS.

Though rare, vitelline duct remnant is an important feature of TARPS, as it can lead to intestinal obstruction. Vitelline duct remnants and persistent left superior vena cava are both due to lack of resolution of relevant embryological features, demonstrating that RBM10 has an important role in this process. Recently RBM10 has been suggested as a susceptibility gene for split hand foot malformation (SFHM) (54). Congenital malformations of hands and feet are indeed a feature of TARPS, ranging from SHFM over missing or abnormal first finger or toes to syndactylies of 2.-3. toes. The phenotypes of individuals with RBM10 variants thus support an association of RBM10 to abnormalities of hands and feet, including HSFM.

All analyzed variants cause some degree of RBM10 LOF. This was reflected in the splicing analysis, which showed a clear pattern of more exon inclusion in response to increased RBM10 LOF. More inclusion correlated with increasing phenotype severity. This was further supported by splicing analysis of selected targets in fibroblasts, which showed larger alterations in fibroblast from an individual with cTARP than in an individual with TARPL. In addition to RBM10s role as a splicing regulator, we hypothesize that it also directly regulates gene expression by acting as a regulator of transcription through the C_2_H_2_-type ZF domain. This is reflected by the larger differences between the number of significantly differentially expressed genes in TARPS than in RAID compared to significant altered splicing events. Thus, the TARPS phenotype observed in individuals with missense variants in the C_2_H_2_-type ZF domain could arise from both altered splicing pattern and transcription regulation. Such dual function of an RBP has previously been described for RBM22, which regulates splicing through its RRMs and gene expression through a ZF-like domain (55).

Whereas the p.L307P missense variant resulted in severely reduced levels of RBM10 protein and caused severe cTARP, other missense variants led to a stable protein and caused a spectrum of milder phenotypes. Functional analysis of three C_2_H_2_-type ZF domain constructs harboring TARPL missense variants showed that the two p.R766 variants had no effect on folding of the domain and were suggested to affect interactions with binding partners of the domain. The p.H784R variant had a larger effect on the Zn^2+^coordination and did also display local misfolding of the domain. Interestingly, the individual with the p.H784R variant (P11) had the most severe phenotype of the three (Supplementary Table S1). The C_2_H_2_-type ZF domain has been shown to be crucial for normal RBM10 protein function and localization in nuclear speckles which is important for splicing regulation (5, 56–58). Recently, an RBM10 somatic C_2_H_2_-type ZF domain variant (p.C761Y) identified in cholangiocarcinoma was shown to decrease binding affinity to SRSF2 (5). Interestingly, we also identified the p.C761Y variant in a TARPL individual (P17) in our cohort.

RBM10 is associated with several spliceosome components localized in splicing speckles (2, 57, 59–61). The C_2_H_2_-type ZF domain is important for RBM10s binding to U2 snRNP in the U2snRNP/branchpoint complex presumably important for splicing regulation (9). Therefore, the missense variants in the C_2_H_2_-type ZF domain, shown to affect the folding or function of the domain, could affect its ability to interact with the U2 snRNP/branchpoint complex and/or affect the localization of the RBM10 protein in the nucleus.

Consequences on domain protein folding of the RRM2 domain constructs correlate with severity of the associated phenotype. The p.L307P and p.H367P domain constructs showed complete misfolding, these were seen in individuals with cTARP or intermediate phenotype, respectively. The p.H367P variant resulted in local misfolding of the RRM2 domain, but did not affect the total amount of RBM10 protein when compared to the two RAID variants p. R332C and p.R332H (Figure 4B), that showed no effect on RRM2 domain folding. The p.R332C and p.R332H variants were shown to have altered RNA-binding affinity.

In RAID individuals the smallest number of changed splicing events, overlapping KO events and genes with altered expression were observed. We hypothesize that altered RNA-affinity modifies the binding of RBM10 to multiple targets, resulting in loss of binding or partial loss of binding to some targets, but also potential binding to new sequence motifs in new target genes. Consistent with this hypothesis, we demonstrated that p.R332C and p.H367P variant proteins had partially or fully lost their ability to regulate *SMN1/2* exon 7 splicing, whereas p.H367P showed GOF towards *ACADM* c.362C>T exon 5 inclusion (Figure 7B). This may indicate that the observed misfolding of the RRM2 domain of the p.H367P variant most likely drastically changes the RNA binding affinity of this protein, resulting in the LOF of splicing regulation of multiple targets and at the same time a GOF splicing regulation of other targets. We hypothesize that this could also be the case for the other RBM10 mutant proteins. In patient and CRISPR samples with the p.R332C variant, we identified splicing changes unique for this variant, reinforcing the hypothesis that the different variants can result in different splicing changes (Supplementary Table S16). Variants altering RNA-specific binding have been observed in other splicing factors as well, including SRSF2 in myelodysplastic syndromes (50).

cTARP syndrome is mainly caused by PTC variants scattered throughout *RBM10,* excluding the alternatively spliced exon 4, and therefore abolish formation of both protein isoforms (RBM10v1 and RBM10v2). Individuals with variants causing PTCs in exon 4, who have the milder TARPL phenotype, were shown to have lower total *RBM10* mRNA levels than controls or other individuals with *RBM10* variants. This is most likely the result of NMD-mediated degradation of the PTC containing exon 4 mRNA transcripts (RBM10v1 isoform), while RBM10v2 mRNA levels are unaffected. An individual with an exon 4 5’splice site variant (P30), resulting in complete exon 4 skipping, thus exclusively expressing a normal version of the RBM10v2 isoform, was shown to have total *RBM10* mRNA amounts comparable to control (Figure 6A). We hypothesize that this individual has a milder phenotype than observed for individuals with PTC variants in exon 4 due to increased amounts of WT RBM10v2. Consistent with this, the individual with the exon 4 5’splice site variant has unaffected splicing patterns of *VCL* exon 19 and *TNRC6A* exon 7, thus indicating that RBM10v2 is able to regulate the splicing of these exons. Furthermore, individuals with PTC variants in exon 4 have lower inclusion of VCL exon 19 and TNRC6A exon 7 than other TARPS individuals (Supplementary Figure S6+7). This supports a model whereby the RBM10v2 isoform is functional.

In conclusion, we demonstrate that the RBM10-phenotypic spectrum consist of TARPS and the new phenotype RAID. We describe a clear genotype-phenotype correlation depending on RBM10 protein LOF, with the largest degree of functional loss correlating the most severe phenotypes. We show that different molecular mechanisms can explain the underlying pathological alterations in RBM10 protein function, and that missense variants in *RBM10* in addition to LOF may also cause GOF effects.

## Methods

Methods for functional studies can be found in Supplementary information.

### Sex as a Biological Variant

Our study exclusively included male individuals because *RBM10* associated syndromes are X-linked recessive.

### Cohort

40 individuals from 32 families were included in the study, including three previously published individuals (15, 31). Of these 40, 16 individuals from 14 families provided blood samples for functional analysis, 2 provided fibroblast, and samples from 8 male relatives were included as healthy control.

### Statistics

For analysis of differential splicing, the default unpaired statistical rMATS model was used for all analyses. For analysis of differential gene expression, read counts were modelled using negative binomial distribution and statistical significance determined with the Wald test. All p-values from RNA-seq data were adjusted to correct for multiple testing using Bejamini-Hochberg method. Quantitative data is represented as means ± standard error of mean (SEM), statistical testing was evaluated using two-sample, two-tailed *Student’s t-test* assuming equal variances

### Study Approval

Approval for studies of families with *RBM10* variants is obtained from The Regional Committee on Health Research Ethics (Project-ID: S-20180082).

RNA-seq data from patients and families will be kept on secure servers due to General Data Protection Regulation (GDPR).

The identification, validation, segregation analysis, and clinical reporting of *RBM10* variants was carried out by each participant’s clinical center. Individuals were identified through the Decipher database (62), Genematcher (63), Matchmaker Exchange (64), and personal network, and invited via their respective clinician to participate in the study. All patients and/or legal guardians were given verbal and written information before signing a written consent for participation in the study. Written consent from patients and/or legal guardians were received for the use of photographs.

## Data Availability

RBM10 CRISPR RNA-seq data is deposited in the ArrayExpress database (https://www.ebi.ac.uk/bi-ostudies/arrayexpress) under accession number E-MTAB-15267. Patient RNA-seq data is not available due to ethical restrictions. The analytical code is available upon request. Values for all presented data are reported in the Supporting Data Values file.

## Supporting information

Supplementary data values

## Data Availability

RBM10 CRISPR RNA-seq data is deposited in the ArrayExpress database (https://www.ebi.ac.uk/bi-ostudies/arrayexpress) under accession number E-MTAB-15267. Patient RNA-seq data is not available due to ethical restrictions. The analytical code is available upon request. Values for all presented data are reported in the Supporting Data Values file.

https://www.ebi.ac.uk/biostudies/arrayexpress

## Author Contributions

First authorship is shared between JMVB and CRF. JMVB has been in charge of the functional studies and is therefore listed first, CRF have conducted the clinical part of the study.

Study concept and design: JMVB, CRF, TKD and BSA.

Generation of experimental data: JMVB, SML, AGM and MMR with contribution from SH, LWS, and NAN.

Analysis and interpretation of experimental data: JMVB, MMR, KB, CRF, TKD, MB, SML, AGM, MS, JV and BSA.

Patient and family recruitment and clinical data analysis: CRF.

Face2gene analysis: CA.

Contribution of clinical data and patient samples: QH, LKH, ML, PS, CS, MS, APAS, HVE, CDL, CVM, AG, DW, JR, JM, VD, KZK, JM, PA, PR, BG, MJMG, JMG, GV, FP, Al, MF, EMM, BK, MDF, TY, KS, AR, AB, GC, ZS, DC, GBF, IV, MCS, BM, LT, CK, OV, AV, MRPB, EC, FBC, LOF, CP, MT, LKW, SFN, MS, VN, VC, KT, VC, JL, VK, ADD.

Manuscript drafting: JMVB prepared first draft with input from CRF, TKD and BSA.

Critical revision of manuscript: JMVB, JV, MS, AGM, SML, CRF, TKD and BSA with input from all authors.

Funding acquisition: BSA.

Study supervision: BSA, CRF.

## Supplementary Material

Supplementary Tables

Supplementary Information including supplementary figures, materials and methods.

Supporting Data Values file

## Acknowledgements

We thank the patients and families for sharing information and contributing to increasing knowledge on this rare disease.

This work was supported by a grant from The Lundbeck Foundation (R286-2018-1739 to JMVB and BSA), Natur og Univers, Det Frie Forskningsråd (0135-00459B and 3103-00329A to BSA), The fund to support clinical research careers in the Region of Southern Denmark (Region Syddanmarks pulje for kliniske forskerkarriereforløb, CFA), and California Center for Rare Diseases within the UCLA Institute for Precision Health (SN, MT, LKW) .

The Euro-MRX project, where the first family had their X-chromosome exome sequencing performed, was financially supported by the EU FP7 project GENCODYS (241995).

Several authors of this publication are members of the European Reference Network on Rare Congenital Malformations and Rare Intellectual Disability ERN-ITHACA. [EU Framework Partnership Agreement ID: 3HP-HP-FPA ERN-01-2016/739516]

This study makes use of data generated by the DECIPHER community. A full list of centers who contributed to the generation of the data is available from https://deciphergenomics.org/about/stats and via email from. DECIPHER is hosted by EMBL-EBI and funding for the DECIPHER project was provided by the Wellcome Trust [WT223718/Z/21/Z].

This study utilized the GeneMatcher platform, a founding member of the Matchmaker Exchange project, to identify potential matches, data shared through the seqr platform (funding provided by National Institutes of Health grants R01HG009141 and UM1HG008900), and use data shared through the PhenomeCentral repository (funded by Genome Canada and Canadian Institute of Health Research).

