## Supplementary data for "Genotype-Phenotype Correlation in RBM10-Associated Syndromes – How Variant Function Shapes a Broad Phenotypic Landscape"

**Supplementary Figures:**

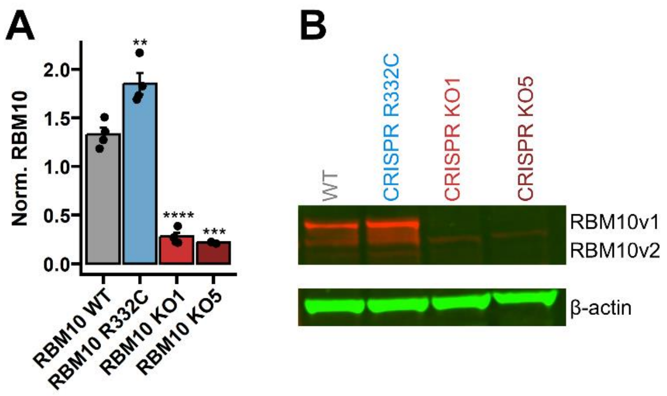


**Supplementary Figure S1** – RPE-1 CRISPR RBM10: **A.** Investigation of RBM10 mRNA expression in WT, RBM10 R332C, RBM10 KO1, and RBM10 KO5. Investigated with RT-qPCR. RBM10 is normalized to RPL13A. n=3. *p<0.05, **p<0.01, ***p<0.001 by two-sample, two-tailed *Student's t-test* assuming equal variances. **B.** Investigation of RBM10 protein expression in WT and CRISPR cell lines by western blotting using fluorescently labelled secondary antibodies.

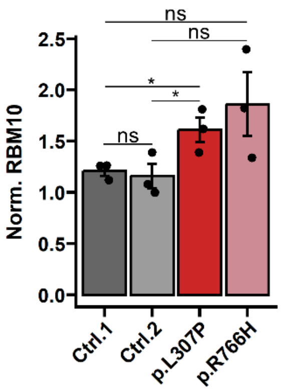


**Supplementary Figure S2** – RBM10 Expression in Patient derived Fibroblast. Investigation of *RBM10* mRNA expression on RNA from patient derived fibroblast from P6 (p.L307P) and P8 (p.R766H) with RT-qPCR RBM10 normalized to RPL13A. *$p<0.05$ by two-sample, two-tailed *Student's t-test* assuming equal variances.

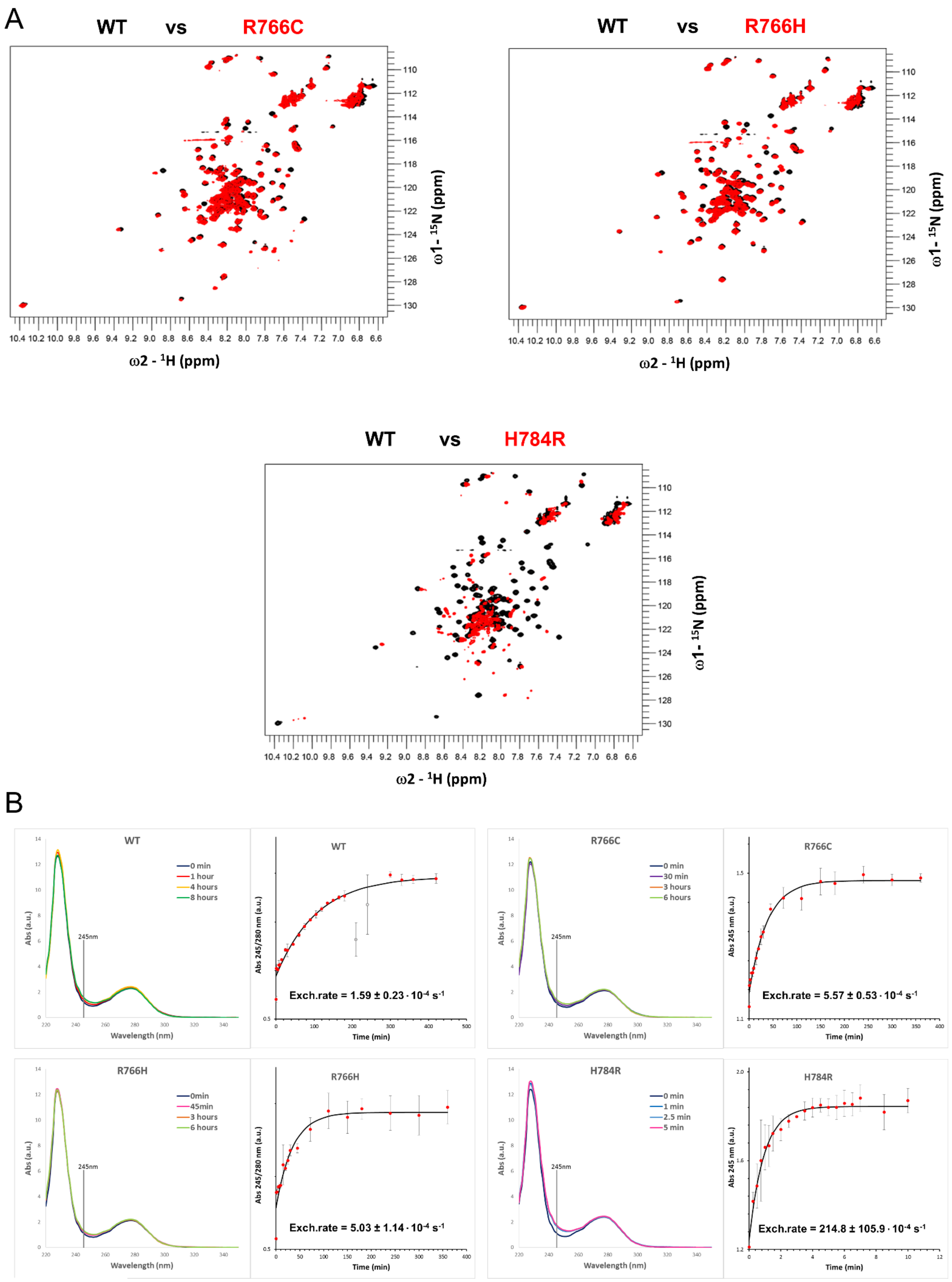


**Supplementary Figure S3 -** C2H2-Type ZF Domain Stability. **A.** 1H-15N HSQC spectra overlay of the wild type RBM10 C2H2-Type ZF domain construct (black) and the different mutants (red): Top left - p.R766C, Top right – p.R766H, Bottom – p.H784R. Both p.R766 mutants show little chemical shift perturbation, suggesting the maintenance of the general fold, while p.H784R mutant presents much more perturbation indicating misfolding of the protein. **B.** $Zn^{2+}$ coordination stability was investigated in mutant C2H2-Type ZF domain constructs and compared to WT C2H2-type ZF domain constructs. The constructs were treated with 5x molar excess of a $Cd^{2+}$-EDTA and the UV-visible absorbance was monitored over time. A band around 245 nm is expected when $Cd^{2+}$ is coordinated. WT exchange rate is slower suggesting a strong $Zn^{2+}$ coordination. R766 mutants show slight decrease in stability with a 2-3X faster exchange. The H784R mutant clearly shows a rapid exchange, which is explained by its unstable coordination.

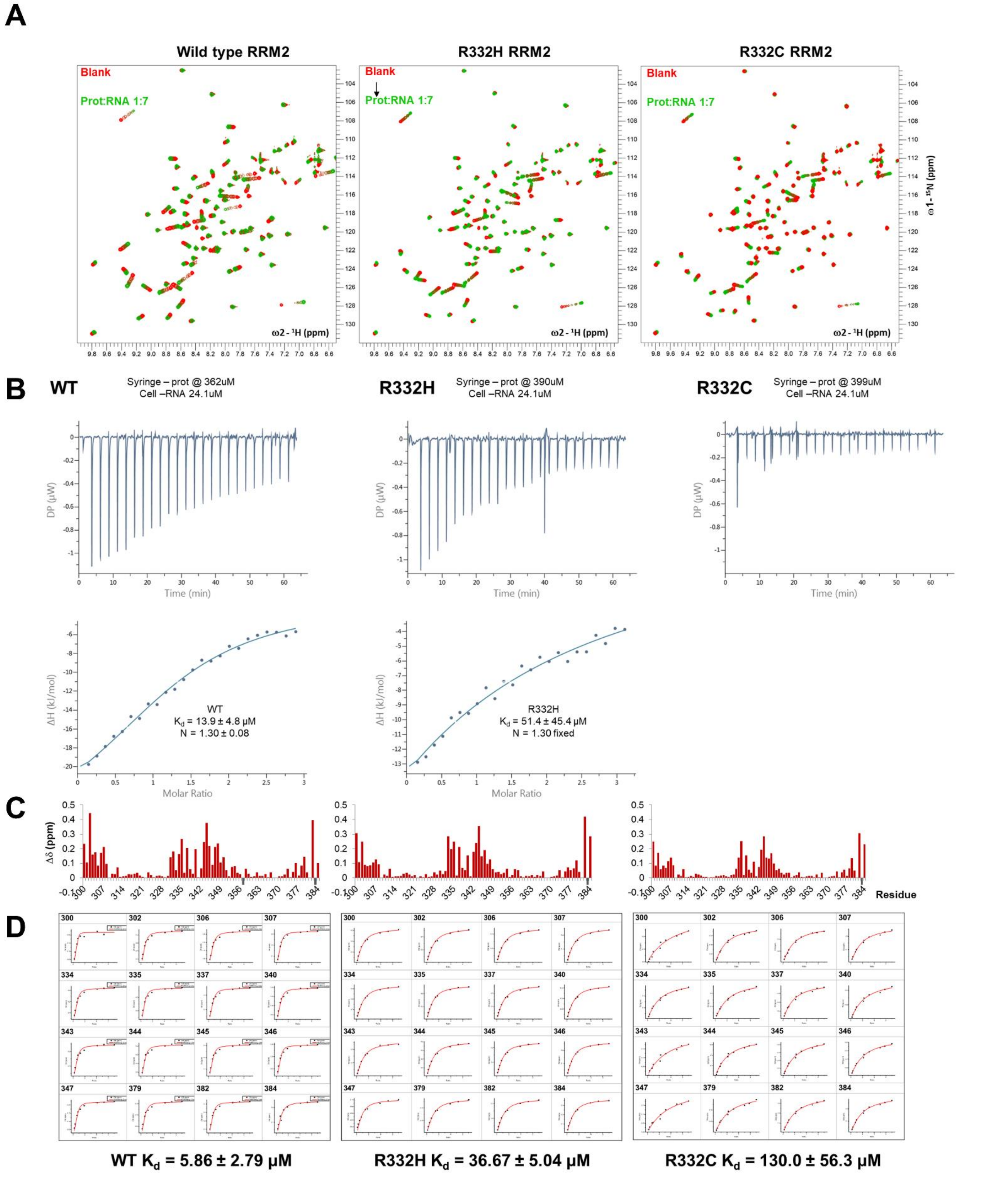


**Supplementary Figure S4 –** p.R332 RRM2 domain binding to RNA. **A.** 1H-15N HSQC spectra acquired for each protein at different titrations points up to 7 molar excess of RNA (red free protein - green 1:7 excess of RNA). **B.** Isothermal titration calorimetry experiments of RBM10 RRM2 constructs (WT, p.R332H, p.R332C) with an RNA oligonucleotide derived from *NUMB* exon 12 (5'-UUGUCUGCUCCC-3'). **C.** Chemical shift perturbation analysis per residue for each titration at the highest RNA concentration (1:7, protein:RNA ratio). **D.** Quantification of the overall affinity for each protein to the RNA oligonucleotide by following the chemical shift perturbation as a function of the protein:RNA molar ratio for 16 selected NH correlation signals presenting highest perturbation.

A

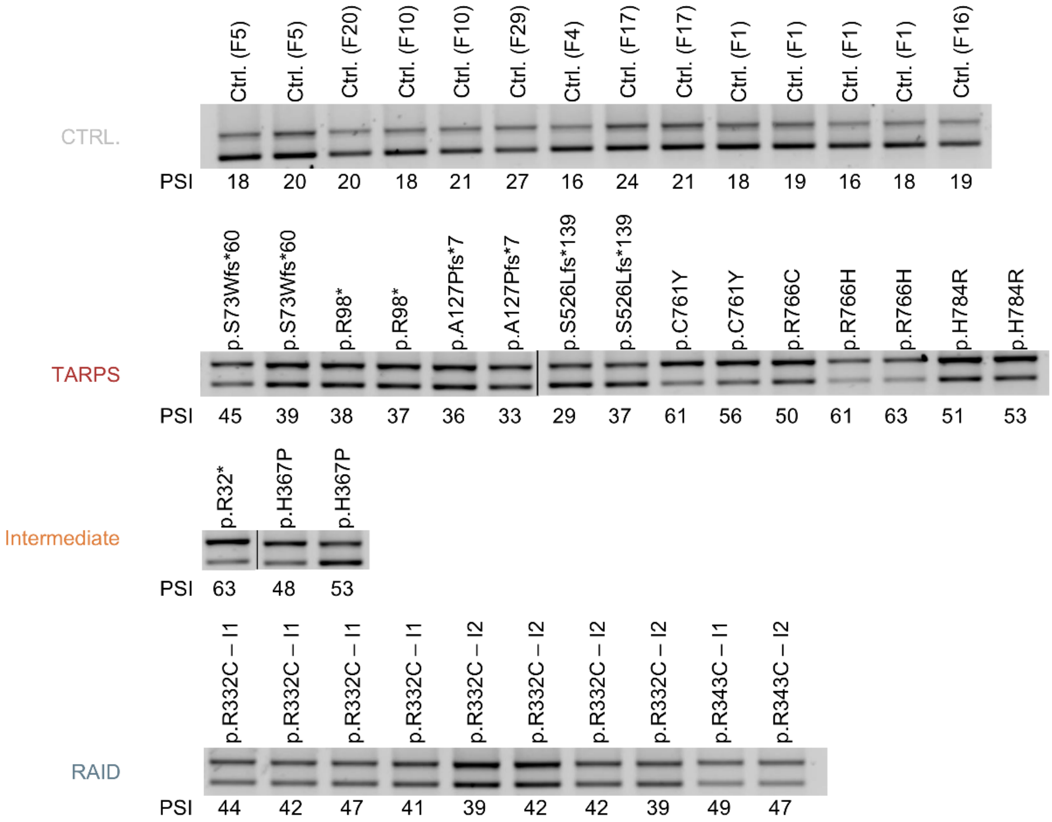


B

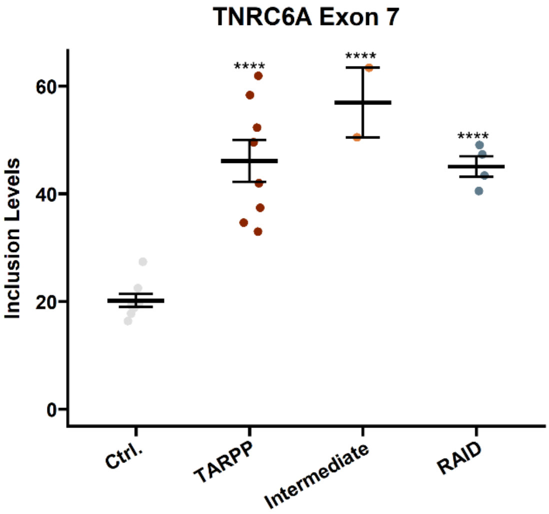


**Supplementary Figure S5** - *TNRC6A* Exon 7 Splicing. **A.** Investigating *TNRC6A* exon 7 splicing in RNA from blood from individuals in the RBM10 phenotypic spectrum compared to healthy males. Individuals are divided into phenotype groups; TARPS (TARP Syndrome), Intermediate and RAID (RBM10 associated intellectual disability). Black line marks non-contiguous gel lanes. **B.** *TNRC6A* exon 7 inclusion levels from **A.** visualized on a jitterplot.

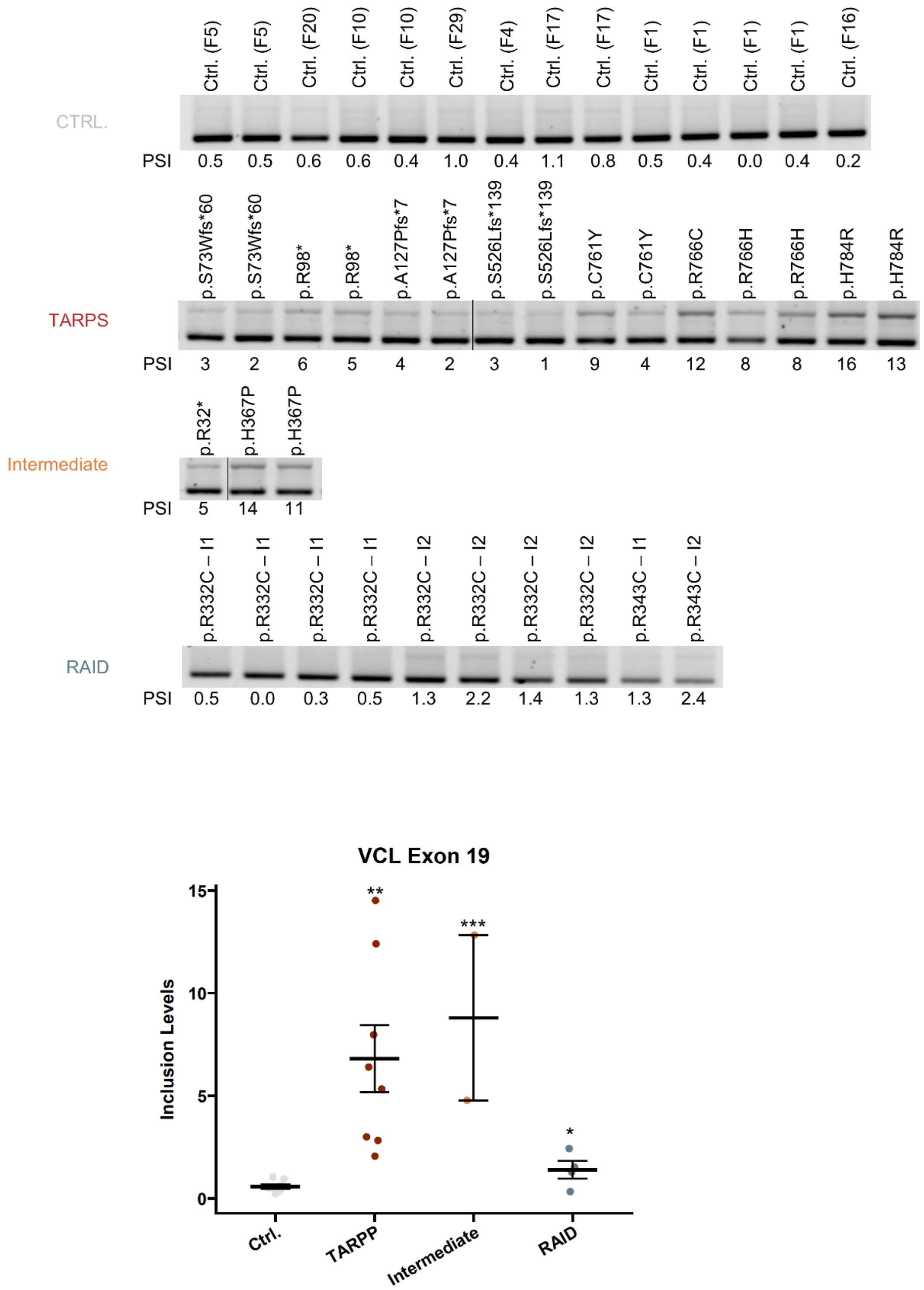


**Supplementary Figure S6** - *VCL* Exon 19 Splicing. **A.** Investigating *VCL* exon 19 splicing in RNA from blood from individuals in the RBM10 phenotypic spectrum compared to healthy males. Individuals are divided into phenotype groups; TARPS (TARP Syndrome), Intermediate and RAID (RBM10 associated intellectual disability). Black line marks non-contiguous gel lanes. **B.** *VCL* exon 19 inclusion levels from **A.** visualized on a jitterplot.

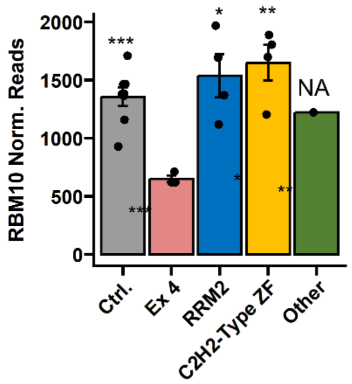


**Supplementary Figure S7** – RBM10 Expression in Patient RNA. Normalized *RBM10* reds from DESeq2 gene expression analysis comparing patients with variants causing STOP in exon 4 (light red, n=3) compared to healthy controls (grey, n=8) and the other individuals included in the study grouped based on location of the variant (RRM2 – n=3, C2H2-Type ZF – n=4 and another variant along the *RBM10* gene n=1). *p<0.05, **p<0.01, ***p<0.001, by two-sample, two-tailed *Student's t-test* assuming equal variances.

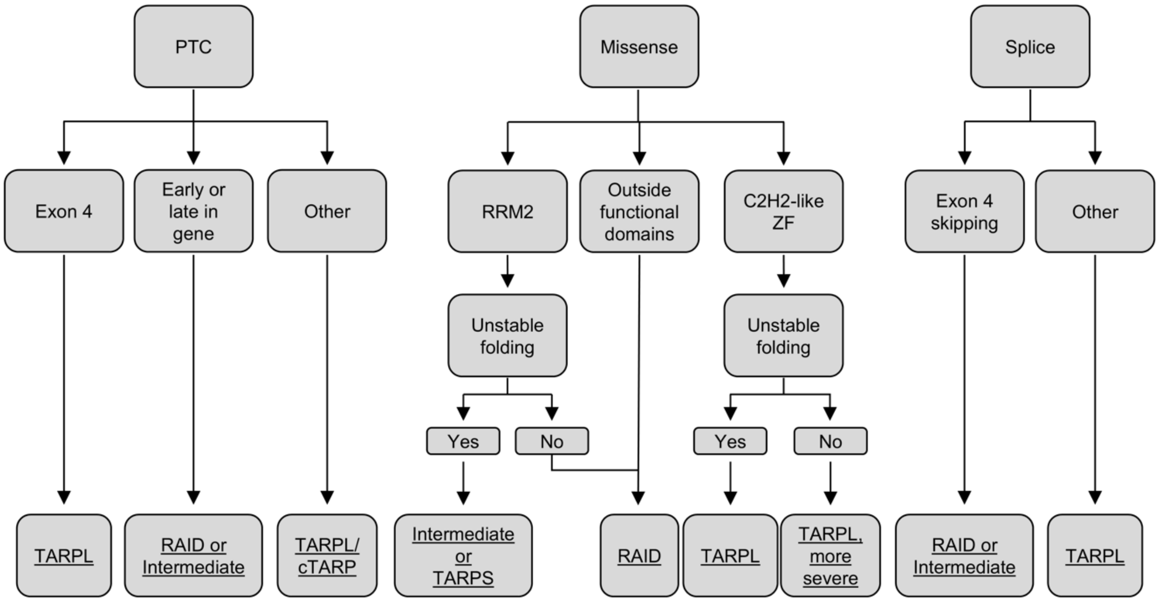


**Supplementary Figure S8** - Experience from present study concerning genotype-phenotype correlation in RBM10. RAID (RBM10 associated intellectual disability), TARPS (TARP syndrome), TARPL (TARP-like), and cTARP (classical TARP).

**Supplementary Methods:**

***RNA Extraction from Blood Samples***

Blood was collected in PAXgene Blood RNA tubes (PreAnalytix, #762165). Samples were left at room temperature for a minimum of 2 h, followed by 24 h at -20 °C, and stored at -80 °C. The samples were transported on dry ice. The RNA extraction was carried out using the PAXgene Blood RNA kit (PreAnalytix, #762174), according to the manufactural protocol. RNA concentration and quality were measured on the Agilent Bioanalyzer 2100 using the Agilent RNA 6000 nano kit (Agilent, #5067-1511).

***Introducing Patient Variants in Human Cell Line using the CRISPR CAS9 System***

RBM10 patient variants were introduced into human hTERT-RPE-1 cells with doxycycline inducible CAS9 (Kind gift from Professor Jens S. Andersens Laboratory, SDU). Cells were stimulated with 1 µg/mL doxycycline 48 h and 24 h prior transfection. RNA-duplexes were formed from crRNA and trancrRNA (crRNA sequences listed in supplementary Table S17) to a final working concentration of 1 µM. Transfection complexes were prepared with RNA duplex, ssDNA donor template, Lipofectamine RNAiMAX (Invitrogen, #13778100) and serum free RPMI media (Sigma, #R0883). hTERT-RPE1 cells were transfected with transfection complex to a final volume of 2.4 mL. Cells were seeded for single cell colonies in a 15 cm culture plate, when single cell colonies were visible by eye, they were transferred to 24-well plates, later split into two wells in 12-well plates. Genomic DNA (gDNA) from CRISPR generated cells were screened for patient variants using Amplification Refractory Mutation System PCR (ARMS-PCR). DNA was extracted from cells by boiling the cell pellet for 15 min in 100 µL 1xTris-HCl pH 7.6, followed by centrifugation for 3 min at 3000 g and collection of the supernatant. 1 uL gDNA was amplified with TEMPase Hot Start 2x Master Mix C (Amplicon, # A230704) using allele specific primers (Primer sequences listed in supplementary Table S18).

***Cell and Transfection***

Patient fibroblasts, control fibroblasts (FB01 and FB07), RPE-1 and CRISPR cells were cultured in RPMI media (Sigma, #R0883) with 10 % Fetal Bovine Serum (FBS) (Sigma, #F7524) and 5 % Pen/Strep (Lonza, #DE17-602F). CRISPR RPE-1 cells were transfected with *RBM10* expression vectors variant 1 (pCMV6-XL4 RBM10, OrigGene, NM_005676) and co-transfected with T7-pCI vector, *SMN[1]* or *ACADM* mini-genes [2] using Xtremegene 9 (Roche, #06365787001), RNA and protein was harvested 48 h after transfection. Cells were frequently tested for mycoplasma throughout the project.

### *RBM10 Protein Analysis*

Western blot analysis was carried out to investigate RBM10 protein levels in human cell lines. Protein was harvested by trypsinizing cells and diluting cell pellets in RIPA Buffer (ThermoFisher Scientific, #89901) supplemented with Halt Proteinase Inhibitor Cocktail (ThermoFisher Scientific, #78430). Protein lysate was treated with Benzonase Nuclease (Merck, #E1014) concentrations were determined by bicinchoninic acid (BCA) assay (ThermoFisher Scientific, #23227). Proteins were separated on a 4-12% Sodium Dodecyl Sulfate – Polyacrylamide gel electrophoresis (SDS-PAGE) (ThermoFisher Scientific, #NP0321BOX) followed by transfer to a PVDF membrane (Cytvia, #15279884). The membrane was probed with an anti-RBM10 antibody (Abcam, # ab224149), anti-T7 antibody (Merck, #69522-4) and anti-β actin (Abcam, #ab8226) used as loading control. Fluorescently labelled secondary antibodies were used (Cytvia, #PA45011 and #PA43009). No-Stain™ Protein Labeling Kit (ThermoFisher Scientific, # A44717) was used to detect total protein amount.

### *RNA Extraction and Analysis*

RNA was harvested from human cell lines using TriReagent (Sigma-Aldrich, #93289) and chloroform extraction. RNA was reverse transcribed into cDNA using High Capacity cDNA reverse transcription kit (Life Technologies, #4368814). The RBM10 expression was analyzed by quantitative PCR using iTaq™ Universal SYBR® Green Supermix (Biorad, #1725125) (Primer sequences listed in supplementary Table S18). RT-PCR was carried out using TEMPase Hot start 2x mastermix C (Amplicon, #A230704) and sequence specific primers (Primer sequences in supplementary Table S18).

### *RNA Sequencing*

RNA from blood samples were prepared for RNA-seq using the NebNext Ultra II Directional RNA library prep kit for Illumina (New England Biolabs, #E7760). Samples were paired-end sequenced (150 bp) using Illuminas NovaSeq 6000.

Raw reads were trimmed for adapter and homopolymers using BBduk2.sh, a part of the BBMap toolset [3], and subsequently aligned with STAR v2.7.5c [4] to the human genome using the hg38 assembly, and known splice junctions from release 106 of the Ensembl gene annotation [5]. All samples were first mapped one time, and novel junctions were then collected from all samples and used to build a new STAR index. All samples were then mapped to this index to produce final alignments. These alignments were used as input to rMATS turbo v4.1.2 [6] in a combined analysis to maintain splice event id numbers across comparisons.

Statistical analyses were then performed in a subsequent step of the rMATS analysis using only the relevant samples, per the rMATS user guide.

For gene expression analyses, we mapped reads to the Ensembl release 106 transcriptome using salmon v1.7.0 [7], and converted transcript estimates into gene counts using tximport v1.24.0 [8]. We then used DESeq2 v1.36.0 [9] to test for differential gene expression between groups, and used apeglm v1.18.0 [10] to adjust log2FC estimates according to the DESeq2 guidelines.

Subsequent comparative analyses of splicing patterns and gene expression were conducted in R v 4.2.1 [11] with volcano plots produced using EnhancedVolcano v1.14.0 [12] and Venn diagrams by VennDiagram v1.7.3 [13]. To produce sashimi-plots, we used an adapted version of ggsashimi [14] that implements calculation of junction means when some samples contain a count of zero for a given junction.

In analysis of blood samples, all available control samples across families were used as control group when examining changes in splicing and gene expression.

***Recombinant Protein Expression and Purification***

DNA sequences encoding RBM10v1 RRM2 (300-384) and C2H2-ZF (726-820) (IDT) were cloned into pET M11 vector using NEBuilder HiFi DNA Assembly Cloning Kit (New England Biolabs). $C_2H_2$-ZF mutants (p.R766H, p.R766C and p.H784R) were prepared in a similar way while RRM2 mutants (p.L307P, p.R332C and p.H367P) were created via site-directed mutagenesis using DNA oligonucleotides (Primer sequences listed in Supplementary Table S18)

Plasmids containing wild type or mutant sequences were transformed into *Escherichia coli* BL21(DE3) cells. Cultures were grown in Luria-Bertani (LB) broth or M9 minimal media supplemented with 1 g/l $^{15}NH_4Cl$ for natural abundant or labelled samples respectively, at 37 ºC until reaching an $OD_{600}$ of around 0.6. Overnight expression at 22 ºC was induced with IPTG (0.5 mM) and then cells were harvested and frozen at -20 ºC. For recombinant expression of $C_2H_2$-type ZF proteins, cultures were supplemented with $ZnCl_2$ (100 µM).

Cell pellets were resuspended in a buffer containing 20 mM Tris pH 8.0, 500 mM NaCl, 10 mM imidazole, 0.5 mM TCEP and protease inhibitors mix (SERVA), and were lysed by cell disruption in a French press. Clear lysates, after centrifugation (at 18,000 g) and filtering, were loaded into Ni-NTA resin. Resin was then washed with 20 mM imidazole containing buffer and the desired protein was eluted with 500 mM imidazole. C2H2-ZF constructs were processed in a similar way but using Zn-NTA resin to avoid cation exchange in the Zn finger. Proteins were simultaneously dialyzed against a buffer without imidazole (20 mM Tris pH 8.0, 500 mM NaCl, and 0.5 mM TCEP) and digested with TEV protease (homemade at a final concentration of around 50 μg/ml). Digested products were loaded in the same NTA resin column to obtain in the flow-through the desired protein, free of protease, tags and uncleaved protein. Finally. a polishing step by size-exclusion chromatography was performed on a HiLoad 16/60 Superdex 75 column (GE Healthcare) equilibrated with 20 mM sodium phosphate, pH 6.5, 100 mM NaCl and 1 mM DTT buffer for the RRM2 constructs or with 10 mM HEPES pH 7.5, 300 mM NaCl and 1 mM DTT buffer for C2H2-ZF versions.

During the lysis of RRM2 p.L307P and p.H367P mutants the proteins heavily precipitated and almost no soluble protein was obtained after the first step of purification. Refolding attempts of the solubilized protein in 8M urea containing buffer by quick buffer exchange in NTA resin or by slow exchange in overnight dialysis were unsuccessful.

Sample purity was checked by SDS-PAGE and protein concentrations were determined from the aromatic contribution to the UV absorbance spectra at 280 nm.

***NMR Spectroscopy***

NMR samples were prepared at concentrations ranging 75-300 μM in buffer containing 20 mM sodium phosphate pH 6.5, 100 mM NaCl, 1 mM DTT, and 10% $D_2O$ for RRM2 proteins or in buffer containing 10 mM HEPES pH 7.5, 300 mM NaCl, 1 mM DTT and 10% $D_2O$ for C2H2-ZF proteins. $^1$H-$^{15}$N HSQC experiments of the different RRM2 and $C_2H_2$-type ZF variants were recorded at 25 ºC with 800- or 600-MHz Bruker Avance NMR spectrometers equipped with cryogenic triple resonance gradient probes. Spectra were processed with NMRpipe (Delaglio et al., 1995) and analyzed using ccpnNMR Analysis software (Vranken et al., 2005).

For RNA titrations, RRM2 wild type, and p.R332C variants at 75 μM were titrated with increasing concentration ratios of *NUMB* derived 12mer RNA oligo (5'-UUGUCUGCUCCC-3') until 7-fold molar excess; $^1$H-$^{15}$N HSQC experiments were acquired at each titration point. The accurate final RNA concentrations were corroborated by measuring the UV absorbance at 260 nm. Assignment of the RRM2 spectra was obtained from BMRB (27055 entry). The chemical shift perturbation (CSP) was weighted between $^1$H and $^{15}$N chemical shifts differences (Δδ) using the following formula (Williamson, 2013):

$$\Delta\delta_{HN} = \sqrt{0.5 \cdot [\Delta\delta_H^2 + (0.14 \cdot \Delta\delta_N^2)]}$$

The CSP data were mapped onto RRM2 structure (PDB 2M2B) using PyMOL software. In addition, CPS values for the 16 amino acids presenting highest perturbation were fitted using Origin2021 (OriginLab) to a single site equimolar binding model described by the following formula:

$$\Delta\delta_{obs} = \frac{\Delta\delta_{max}}{2}\left(\left(1 + ratio + {K_d}/{P_0}\right) - \sqrt{\left(1 + ratio + {K_d}/{P_0}\right)^2 - 4ratio}\right)$$

With $\Delta\delta_{max}$ corresponding to the chemical shift perturbation at saturation of binding, *ratio* the molar ratio of protein and RNA, $K_d$ the dissociation constant and $P_0$ the protein concentration, which was assumed to be constant for all the titration points.

***Isothermal Titration Calorimetry (ITC)***

Experiments were carried out on a MicroCAl PEAQ-ITC (Malvern Instruments, UK) at 25ºC in 20 mM sodium phosphate pH 6.5 and 100 mM NaCl. RRM2 variants (350-400 µM) were titrated into a solution of the NUMB derived 12mer RNA (24 µM). Experiments were performed in duplicate with injections of 1.5 µl (0.4 µl for first point) separated by 150 s delays to recover thermal power baseline and continuous stirring in the cell (750 rpm) for correct mixing. The reference cell was filled with water in all experiments. Data were fitted to one-site binding model with Malvern PEAQ-ITC Analysis software (Malvern Instruments).
